# Transcranial Focused Ultrasound Alters Conflict and Emotional Processing, Physiology, and Performance I: Dorsal Anterior Cingulate Cortex Targeting

**DOI:** 10.1101/2020.11.25.20234401

**Authors:** Maria Fini, William J. Tyler

## Abstract

The dorsal anterior cingulate cortex (dACC) operates as an integrator of bottom-up and top-down signals and is implicated in both cognitive control and emotional processing. The dACC is believed to be causally involved in switching between attention networks, and previous work has linked it to cognitive performance, concentration, relaxation, and emotional distraction. The present study was designed to evaluate the feasibility of influencing default mode network (DMN) activity and emotional attention by targeting and modulating the dACC with transcranial focused ultrasound (tFUS). Subjects were divided into two groups, one receiving MR-neuronavigated tFUS to the dACC and the other an identical, but inactive tFUS sham. Subjects performed a modified version of the Erikson flanker paradigm using fear and neutral faces as emotional background distractors. Our observations demonstrate that tFUS can be targeted to the human dACC to produce effects consistent with those expected from relaxed contention, including significantly reduced reaction time slowing due to emotional distractors, and an increase in parasympathetic markers of the HRV. These results suggest that tFUS altered emotional processing and enhanced sustained attention, perhaps by facilitating reduced attentional engagement with emotional distractors and reduced need for attention switching evidenced by significant effects on event related potentials (ERPs), reduced alpha suppression, and modulation of delta and theta EEG activity. We conclude that the dACC represents a viable neuroanatomical target for tFUS in order to modulate DMN activity, including emotional attention, conflict resolution, and cognitive control. These effects of dACC-targeted tFUS may prove useful for treating certain mental health disorders known to involve perturbed DMN activity, such as depression and anxiety.

## INTRODUCTION

Executive attention is widely studied and incredibly important in many life functions for survival, but also plays a key role in emotional wellbeing. There is a growing body of evidence that a few structures forming the cingulo-opercular network are critical to establishing and maintaining executive attention (Sadaghiani and D’Esposito, 2015). The network demonstrates demand-modulated activity in a broad range of cognitive tasks including spatial attention (Eckert et al., 2009), sustained focus (Dosenbach et al., 2007), and meditation (Hölzel et al., 2007). The dorsal anterior cingulated cortex (a major hub in the cingulo-opercular network) is crucial in cognitive behavioral performance as well as emotional regulation (Bush et al., 2000; Posner et al., 2019), and thought to monitor and resolve conflict and action outcome (Botvinick, 2007; Dosenbach et al., 2007). The dACC is both anatomically and functionally interposed between the default mode network (DMN) and anti-correlated dorsal attention network (DAN) (Fox et al., 2005; Fox et al., 2009). Activation of the DMN is associated with mind wandering and self-referential thoughts, including negative rumination (Andrews-Hanna et al., 2014; Mason et al., 2007); it is inversely correlated with performance (Drummond et al., 2005; Polli et al., 2005), and is suppressed during cognitive tasks (Raichle et al., 2001). The dACC may execute is role in executive control by flexibly coupling with either network (Spreng et al., 2012; Sridharan et al., 2008; Vincent et al., 2008). Additionally, these regions are central to both the dorsal (goal directed behavior) and ventral (saliency) attention systems (Dosenbach et al., 2006), and act as a hub between the two (Eckert et al., 2009; Seeley et al., 2007). It has been suggested that the dACC plays a key role improving recognition and resolving a conscious percept while dealing with distraction (Hampshire et al., 2010; Vaden et al., 2013).

The dACC receives both top down and bottom-up input from cortical and subcortical structures and is highly connected to the prefrontal cortex, striatum, hippocampus, and the amygdala (Beckmann et al., 2009; Cassell and Wright, 1986; Rushworth et al., 2007). In addition to its role in deciphering conflict, effortful perception (Wild et al., 2012), and alertness (Coste and Kleinschmidt, 2016), the dACC plays a role in mitigating emotional distraction (Iannaccone et al., 2015; Iordan et al., 2013; Shafer et al., 2012). The dACC demonstrates a clear functional overlap between error processing, pain (emotional suffering), and cognitive control (Albert et al., 2010; Cavanagh and Shackman, 2015; Egner et al., 2007; Foland-Ross et al., 2013; Haas et al., 2006; Kanske et al., 2012; Lane et al., 1998; McRae et al., 2008; Shafer et al., 2012; Shafritz et al., 2006; Wang et al., 2008; Wessel et al., 2012; Whalen et al., 1998; Yang et al., 2014). Activity in the dACC is correlated with emotional awareness (McRae et al., 2008), and changes in the dACC reflect alterations in the broader conscious experience (Aftanas and Golocheikine, 2001). Grey matter thickening is seen in the dACC in both reactively short (Tang et al., 2010), and long-term meditation training; conversely, this anatomical area exhibits cortical thinning in individuals with ADHD (Grant et al., 2013). Chronic pain disorders are known to be correlated with attentional deficits in both humans (Dick and Rashiq, 2007) and animals (Rochais et al., 2016) and linked to the dACC. For example, in patients with chronic low back pain there is a significantly lower engagement of the dACC during cognitive interference (Mao et al., 2014). Emotion can also interfere with attention as a form of distraction as evidenced by attentional biasing towards negative emotions in anxiety disorders (Mogg and Bradley, 2016). Also, shifts in attention and inability to sustain focus and are linked to anxiety and depression (Ólafsson et al., 2011).

There is a clear bidirectional link between emotion and executive function (Inzlicht et al., 2015; Lindström and Bohlin, 2011; Okon-Singer et al., 2015; Sarapas et al., 2017), which has larger implications in human experience and wellbeing. It is hypothesized that the above relationship makes the dACC an extremely promising anatomical target for neuromodulation to improve task performance in during conflict and emotional distraction. We further hypothesize that stimulating this area may also modulate emotional affect and physiological response to fearful faces and increased cognitive load.

Although numerous brain stimulation modalities exist, transcranial focused ultrasound (tFUS) has been of specific interest for its potential to modulate cognition (Fini and Tyler, 2017). Non-thermal, low-intensity, pulsed tFUS has been shown to induce changes in EEG (Legon et al., 2014) and fMRI (Kim et al., 2017; Lee et al., 2016a), as well as perceptual (Lee et al., 2016a; Sanguinetti et al., 2016) and mood changes (Sanguinetti et al., 2020). Unlike any other noninvasive neurostimulation technique, tFUS has a high spatial resolution (on the order of millimeters) and can penetrate deep into the brain (Bystritsky and Korb, 2015).

To test our the above hypothesizes, tFUS was delivered in a trial-by-trial fashion to the dACC while subjects were performing a modified version of the Erickson Flanker task (Eriksen and Eriksen, 1974) in which emotional faces (either fear, neutral, or scrambled) were displayed in the background. The Flanker task is often used as a measure of cognitive control and response to interference. Subjects were asked to report the direction of a middle arrow flanked by two arrows on either side: pointing in the same direction (congruent: > > > > >) or opposite direction (incongruent: < < > < <). The task produces well-defined electroencephalographic (EEG) components and error responses. EEG and heart rate changes were recorded. Performance was measured by reaction times, accuracy, and conflict adaption. In previous studies, EEG frontocentral theta and delta activity can be seen during conflict processing, and post-error (Debener et al., 2005; Iannaccone et al., 2015). Preceding trials with emotional faces induces response slowing and recruits the cingulo-opercular network (Papazacharias et al., 2015). Although emotion cannot be measured directly, survey was collected data using the Positive and Negative Affect Scale (PANAS) (Crawford and Henry, 2004). If successful in improving performance or mood, neuromodulation has broad implications for reducing the susceptibility to distraction in healthy individuals for use with meditation, as well as treating the clinical symptoms of ADHD, depression, and anxiety.

## RESULTS

Subjects were asked to perform a modified version of the Erikson flanker task (Eriksen and Eriksen, 1974) in which emotional faces (fear, neutral, or scrambled) were presented as distractors behind the flanker arrows (**Figure 1A**). Twenty-eight healthy volunteers were divided into two groups: one receiving sham stimulation (Sham group), and the other receiving active tFUS to the dACC. Sham and stimulation began 28 ms prior to the onset of the faces and distractor arrows. Each experiment session began and ended with 100 simple flanker trials (baseline, post-stimulation) where white arrows were presented on a black background, and both groups received sham (see Methods). In the stimulation group, MRI-guided neuronavigation was used to target tFUS to the dACC during the main trials. A mean stimulation location of x = 2.9 ± 0.8, y = 22.2 ± 1.7, z = 32.8 ± 1.6 (mean ± SEM) was recorded (Figure 1B-D).

**Figure 1:**
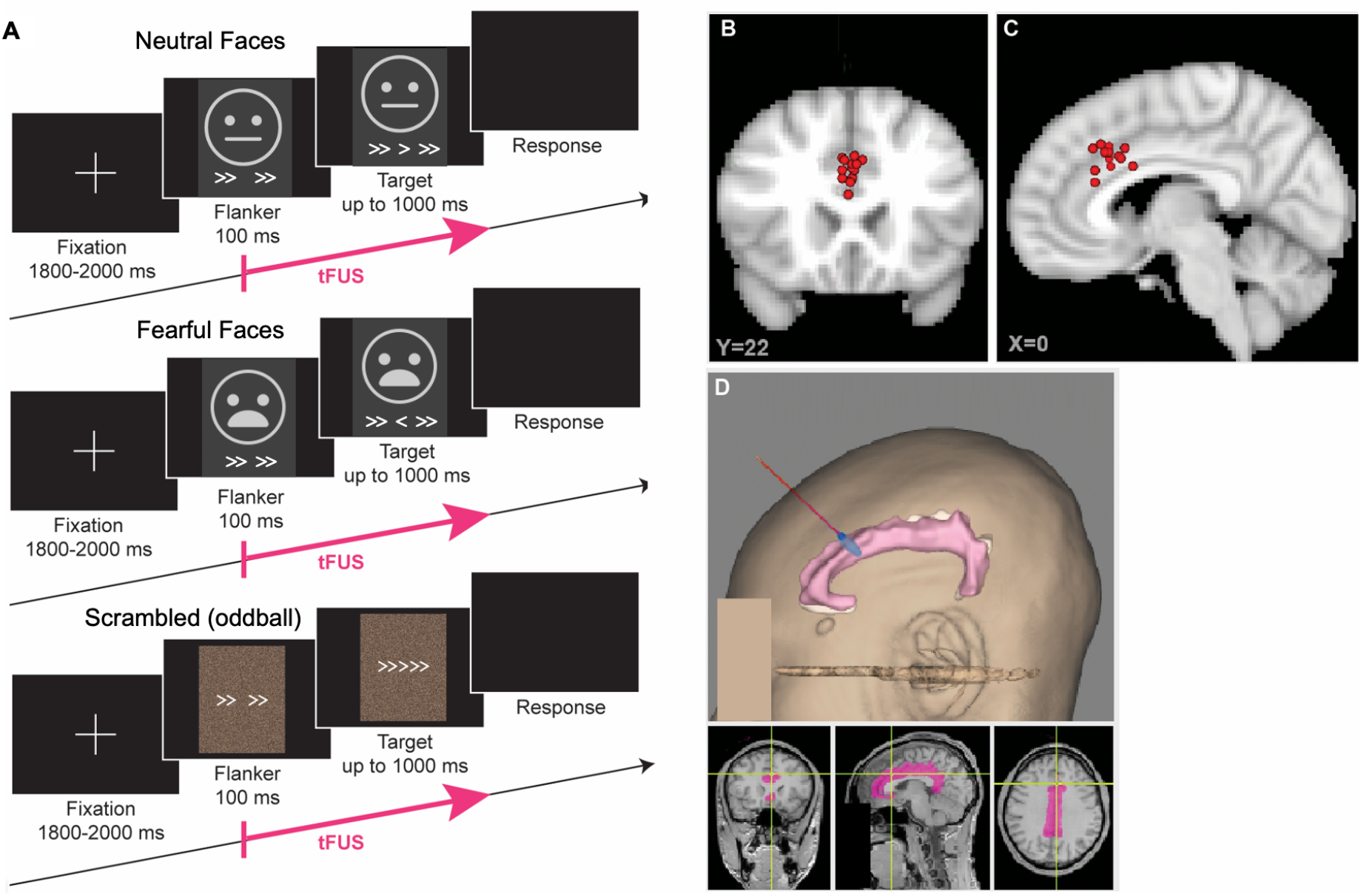
Experimental protocol and stimulation locations. (**A**) All main trials utilized the protocol above. Trials consisted of neutral (top), fearful (middle), or scrambled (bottom) faces. The fearful and neutral faces appeared with equal frequency, while the scrambled trials occurred only 1/50 as an oddball. The stimulation group received online tFUS in all main trials. Stimulation began 28 ms prior to the onset of the distracter flanker arrows and the face image, and lasted 500ms. Both groups were presented with a sham sound during this time period. (**B-C**) Stimulation locations of all subjects mapped onto the MNI brain in coronal (y = 22) and sagittal (x = 0) planes. (**D**) An example of neuronavigation in a single subject. Upper panel shows a 3D reconstruction of the subject’s head, with the cingulate cortex highlighted in pink, beam trajectory in red, and estimated beam focus in blue. Lower panel shows subject’s structural MRI with the cingulate cortex highlighted. Estimated tFUS beam focus center is at the crossing of the green lines.

### Influence of dACC tFUS on target-locked ERPs

Each time point in the ERP response at FCz was subjected to permutation testing across groups (Figure 2A, scalp map details can be seen in Figure S1, alone with ERPs at parietal P4). Results showed that in both the fear and the neutral condition, the distractor elicited D-N1 (first frontocentral negative peak following presentation of distractor face and flanker arrows) shows both a larger negative peak and an earlier onset in the group receiving tFUS to the dACC. Significant differences start as early as 60 ms in the neural condition, and 68 ms in the fear condition. The Sham group shows a longer D-P1 (first frontocentral positivity) than the tFUS group, which is significant in the neutral congruent trials. Additionally P3 has an earlier onset in the Sham group than tFUS in neutral incongruent trials.

**Figure 2.**
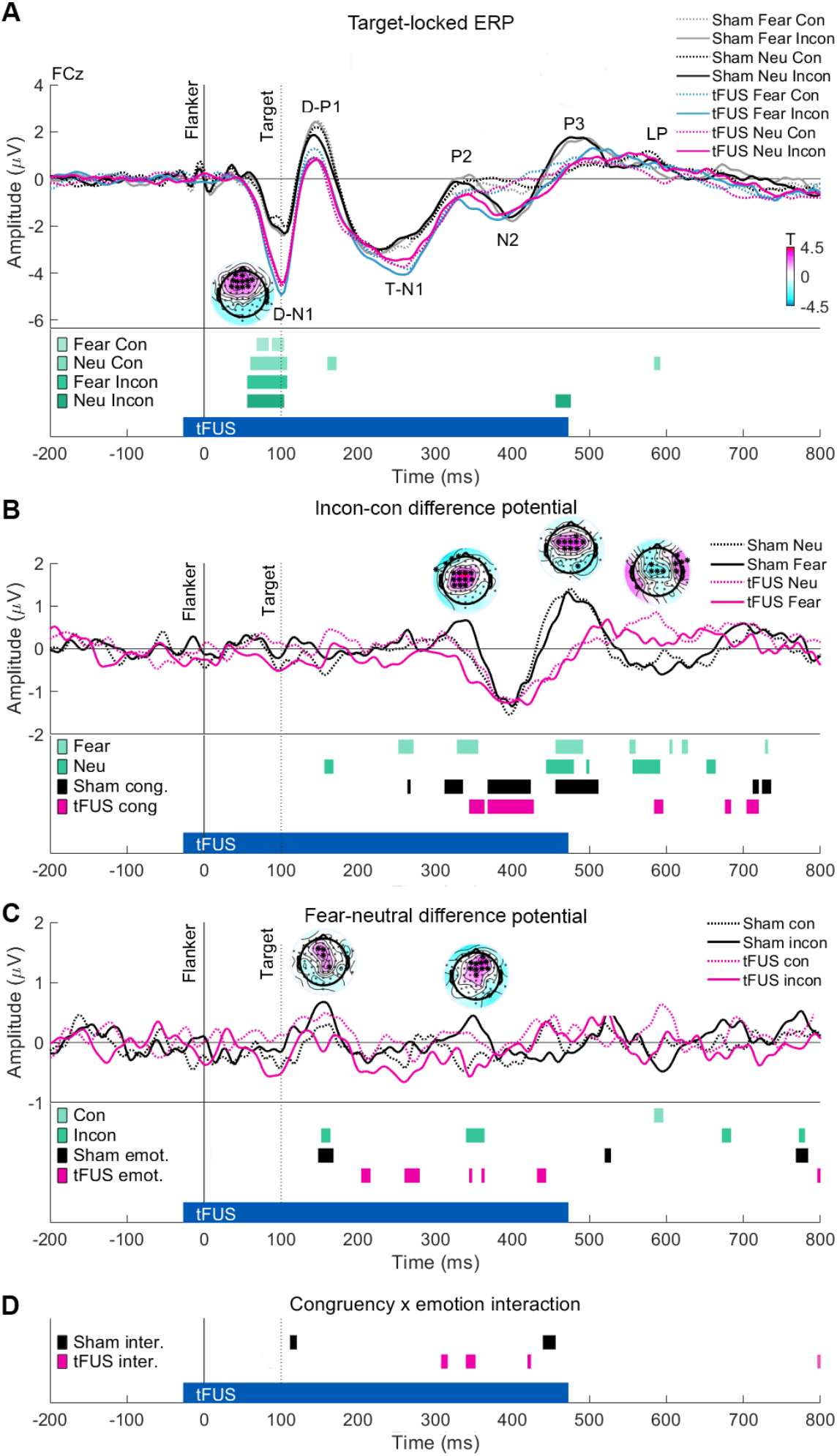
Target-locked event-related potentials at FCz. (**A**) ERP at FCz to neutral and fear trials in both congruency conditions. Time 0 ms marks the presentation of the faces and distractor arrows; time 100ms marks the onset of the target arrow. The tFUS stimulation period is marked with a blue bar at the bottom figure (starts 28 ms prior to face presentation and lasts 500 ms). ERP peaks are labeled. Scalp maps represent T values from permutation testing across groups at 96 ms in the fear incongruent condition, scale on bottom right; positive values indicate tFUS more negative than Sham, negative values: tFUS more positive than Sham. Asterisk represents significant electrodes (*p < 0.05). (**B**) Linear subtraction of incongruent from congruent trials to produce an incon – con difference potential. Lower panel displays significant differences across groups for each emotion condition (fear, neutral), and significant congruency effects for each group (RM-ANVOA). Scalp maps represent T values for permutation testing across groups in the fear condition at P2, P3, and LP. (**C**) Linear subtraction of neutral from fear to create fear - neutral difference potential. Lower panel displays significant differences across groups for each congruency condition (con, incon), as well as significant emotion effects for each group. Scalp T-maps displayed for D-P1 and P2. (**D**) Congruency × emotion interaction effect for each group. Abbreviations: con: congruent, incon: incongruent. Related to Figure S1, Table S2, and Table S3. See also Figure 1.

Individual peak-to-peak amplitude and latencies were assessed at FCz using permutation testing and confirmed this result (Table S1, S3). There is greater amplitude a D-N1 in all conditions in the tFUS group (fear congruent: p = 0.013, neutral congruent: p = 0.019, fear incongruent: p = 0.017, neutral incongruent: p = 0.047). No other significant differences were found across group for ERP peak-to-peak amplitude (all p > 0.15, Table S1).

Similarly, peak latency was compared across groups. In both congruent fear and neutral conditions there was an earlier P3 peak in Sham (fear: 476 ± 5 ms, neutral: 476 ± 5 ms) compared with tFUS (fear: 500 ± 4 ms, neutral: 500 ± 4 ms) groups (fear: p = 0.009, neutral p =0.009). No other latency effects were found (all p > 0.08, Table S3).

### Influence of tFUS targeted to the dACC on incongruent – congruent difference potentials

Congruent potential was subtracted from incongruent potential to create an incongruent – congruent (incon – con) difference potential (Figure 2B). Permutation testing was used to compare each face type across groups and RM-ANOVA was performed within groups [2 congruency conditions (con, incon) × 2 face types (neutral, fear)]. In the neutral condition, there are significant differences across groups in the time range of P3 and the late potential (LP), while the fear condition; there are significant differences across groups in the time ranges of T-N1, P2, P3, and LP. Both groups exhibit a significant congruency effect at N2 (peak more negative in incongruent than congruent), but only in the Sham group is there a significant congruency effect at P3. Indeed, there is markedly smaller incon – con frontocentral positivity at P3 for tFUS group compared with Sham. Additionally, P3 is slightly faster in sham neutral compared fear trials, and significant congruency × emotion interaction can be seen here. At LP incon – con potential is more positive in the tFUS group than Sham; a significant main effect of congruency is observed in the tFUS group during this time period and significant group difference (primarily on neutral trails). At the earlier P2 peak, the Sham group exhibits a significant effect of congruency. In the fear condition, incongruent P2 is more positive than congruent in the Sham group, but this is not the case the tFUS group, and a significant difference across groups can be seen.

Additionally, the tFUS group exhibits a congruency × emotion interaction in this time range between P2 and N2 peaks indicating that N2 onset latency is earlier in the fear condition.

### Influence of tFUS on fear - neutral difference potentials

A linear subtraction of neutral from fear trials was performed to produce fear – neutral difference potential (Figure 2C). A significantly more positive frontocentral D-P1 is seen in the Sham group for fear than neutral trials (a significant main effect of emotion is observed here), yet this is not the case for the tFUS group. Indeed on incongruent trials, a significant difference across groups is seen at frontocentral electrodes. The tFUS group does exhibit a significant effect of emotion at T-N1 (enhanced negativity for fear compared with neutral), and there is a significant difference across groups in the incongruent condition at centro-parietal electrodes (Figure S1). Furthermore, there is a significant difference across groups in incongruent trials at P2 across frontocentral electrodes further supporting the finding P2 and therefore N2 onset is earlier in the tFUS than Sham on fear trials.

### Effects of dACC tFUS on within-group ERP amplitudes

Testing peak-to-peak amplitudes within subjects across all congruency and emotion conditions with nonparametric Friedman’s test further support findings described above (Table S2). Results showed D-N1 – D-P1 amplitude was significantly different across conditions in the Sham group (χ^2^(3) = 7.63, p = 0.048) but not the tFUS group (χ^2^(3) = 3.74, p = 0.29). However, the following D-P1 – TN1 was statically different across conditions not in the Sham group (χ^2^(3) = 5.06, p = 0.089), but in tFUS group (χ^2^(3) = 10.85, p = 0.013), with substantial difference across fear and neutral congruent trials (p = 0.09), and significant differences between fear congruent and neutral incongruent trials (p = 0.034). Both groups exhibited a significant difference between conditions at P2 – N2 (Sham: χ^2^(3) = 30.43, p <0.001, tFUS: χ^2^(3) = 19.06, p <0.001) and N2 – P3 (Sham: χ^2^(3) = 31.89, p <0.001, tFUS: χ^2^(3) = 17.03, p = 0.001) where differences were seen between congruency conditions. Post-hoc statistics show at P2 – N2, there are significant differences between congruent and incongruent trials in both groups for neutral trials (Sham: p = 0.009, tFUS: p = 0.003), but in fear condition, this was only statically significant after Bonferroni correction in the Sham group (Sham: p < 0.001, tFUS: p = 0.059). Both groups showed a significant difference between neutral congruent and fear incongruent (Sham: p < 0.001, tFUS: p = 0.005).

### Effects of dACC tFUS on oddball trials

Analysis of ERP on oddball trials demonstrates that similar to the other trial conditions, there is a larger initial frontal negative deflection in tFUS group compared with sham (Figure S2). Additionally, on incongruent oddball trials, the tFUS group demonstrates both an earlier onset, and a more robust peak at 400ms (N2).

### Parietal time-frequency response suggests tFUS to the dACC affects processing at multiple frequency bands

Event-related spectral perturbation (ERSP) data was calculated and compared at parietal (P3/P4) and frontocentral (FCz) across groups with permutation testing and RM-ANOVA, as well as within groups (RM-ANOVA). All data is reflected as dB increase from baseline (200 ms pre-stimulus baseline used). Comparing groups at parietal (ERSP data pooled across P3 and P4) electrodes reveals significant differences in the delta, theta, and beta bands (Figure 3). There were no group differences at the initial event-locked theta peak (4 – 8 Hz). However, the Sham group exhibits post trial theta suppression in all conditions but greatest in the fear condition, and a significant main effect of emotion was found here for the Sham group (Figure 3C). The tFUS group, in contrast, did not show any theta suppression, but rather a second peak of theta activation in frontal and parietal-occipital electrodes in all conditions peaking between 1000 – 2000 ms post stimulus.

**Figure 3.**
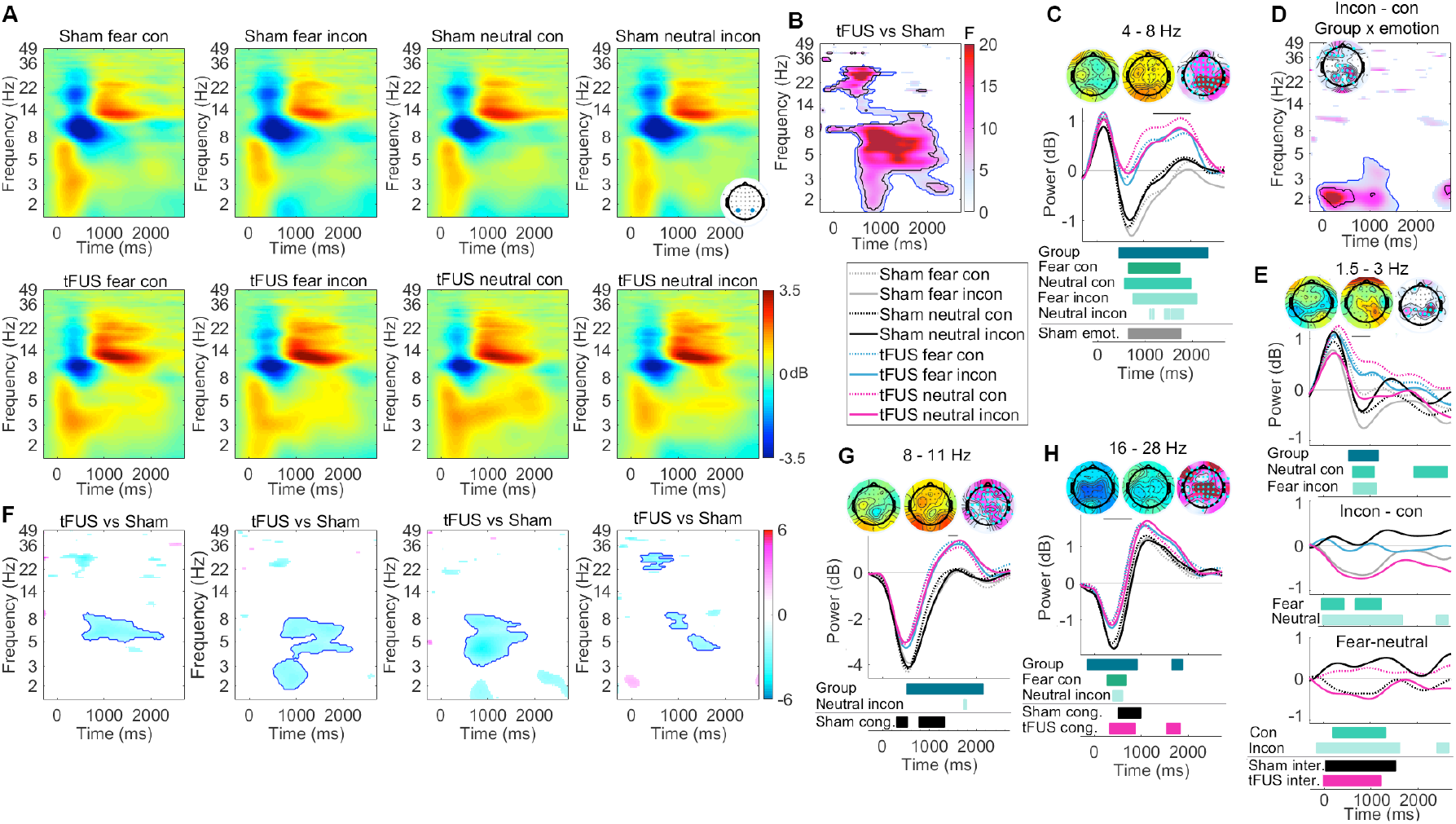
Target-locked time-frequency response at parietal electrodes. (**A**) ERSP data time-locked to target presentation (flanker and faces at 0 ms, target at 100 ms) averaged across P3/P4 (data displayed as dB change from baseline). (**B**) Results of two-way RM-ANOVA (main effect group). In **B, D**, and **F** only significant (p < 0.05) statistical data is displayed; blue lines represent threshold-based clustering; black lines represent statistical significance after FDR correction. (**C, G, H**) Power over time in various frequency bands. Theta (4-8 Hz) displayed in **C**, alpha (8-11 Hz) in G, and beta (16-28 Hz) in H. Bars below represent significant differences across groups using RM-ANOVA (Group) and permutation testing for each condition (green: fear con, fear incon, neutral con, neutral incon), as well as main effect of congruency (cong.) and emotion (emot.) for each group (RM-ANOVA within groups). Scalp maps represent power for time period indicated by horizontal bar above data (left sham, middle tFUS, right **F** values for main effect of group with scale as in B, electrodes significant after FDR correction indicated with cyan dot. (**D**) Interaction effect of group × emotion for incongruent – congruent contrast power. Scalp map displayed for peak significance (0 - 500 ms, 1.5 - 3 Hz). Scale same as **B**. (**E**) Expansion of delta effects displayed in **D**. Top plot shows power over time and scalp maps as in **C, G**, and **H**. Middle plot shows incon – con contrast power for which data is calculated in **D**. Plot line colors are consistent with legend for **C** (gray: Sham fear, black: Sham neutral, blue: tFUS fear, magenta: tFUS neutral). Bars below plot represent significant differences across groups (permutation testing) in incon – con contrast data for each emotion condition (fear, neutral). Bottom plot shows fear – neutral contrast power. Bars below represent significant differences across groups for incongruent (dotted lines) and congruent (solid lines) conditions. Under this, significant congruency × emotion interactions are displayed for each group (inter.). (**F**) Displays T-values from permutation testing across groups for each condition displaced in **A**.

In the alpha range (8 – 11 Hz), the Sham group had larger and longer lasting event-locked alpha suppression than the tFUS group. In the Sham group, this suppression contributed to congruency processing, as there was a significant main effect of contingency. Following the alpha suppression, the tFUS but not the sham group showed post suppression alpha activation largest in parietal and occipital electrodes, but also present in frontocentral electrodes. Permutation testing revealed a significant difference across groups on neutral incongruent trials. This effect was significant over frontal, central, right parietal and occipital electrodes.

In beta-band (16 - 28 Hz), both groups exhibited event-locked beta suppression in parietal electrodes, but this was of significantly larger amplitude in the Sham group compared with tFUS in parietal, central, and frontal electrodes. Permutation testing showed significant differences in the fear congruent and neutral incongruent conditions. Both groups exhibited significant congruency effects during this beta suppression.

In the delta-band (1.5 – 3 Hz), both the Sham and tFUS groups exhibited significant congruency × emotion interaction effects over the first 1000 ms of the epoch, and there was a significant group × emotion interaction effect at parietal and frontal electrodes in incon-con contrast data. Assessing delta activity along with incon – con contrast power and fear – neutral contrast power unpacks this finding (Figure 3E). The tFUS group showed significantly longer target induced delta activation than the Sham group in neutral congruent and fear incongruent trials. Assessing incon-con contrast showed that in the fear condition, the tFUS group showed no differences in delta between incongruent and congruent trials; in both conditions delta power increased and then slowing returned to baseline over the trial epoch. However, in the Sham group, following initial activation, power quickly returned to baseline on congruent trials and showed a small suppression on incongruent trials, producing a negative incon-con contrast. On neutral trials, following the initial delta activation, both congruent and incongruent Sham, and incongruent tFUS delta returned to baseline around 800 ms post-stimulus, but on neutral congruent trials, delta power remained above baseline for 2000 ms in the tFUS group. Therefore, on neutral trials, incon-con power was negative in the tFUS group, and around zero in the Sham group. Indeed, comparing both fear and neutral incon – con power across groups yielded significance in both fear and neutral conditions. Furthermore, fear – neutral power was negative in tFUS incongruent and Sham congruent trials, while being slightly positive in tFUS congruent and Sham incongruent trials. Fear – neutral power significantly differed across groups in both congruency conditions.

### Frontocentral time-frequency response and congruency effects produced dACC tFUS

ERSP data at frontocentral FCz showed significant differences between groups in the delta, theta alpha, and beta range (Figure 4). In delta range, the tFUS group had an earlier onset of event-locked delta (permutation testing showed significance on neutral congruent trials). Both groups showed small but significant congruency effects in the delta range, this effect is earlier and larger in sham group on neutral trials compared with fear trials (significant difference between groups for incon – con contrast power; significant congruency × emotion interaction in the Sham group). Additionally, the Sham group showed an emotion effect beginning at 1000 ms due to suppression of delta power on fear incongruent trials.

**Figure 4.**
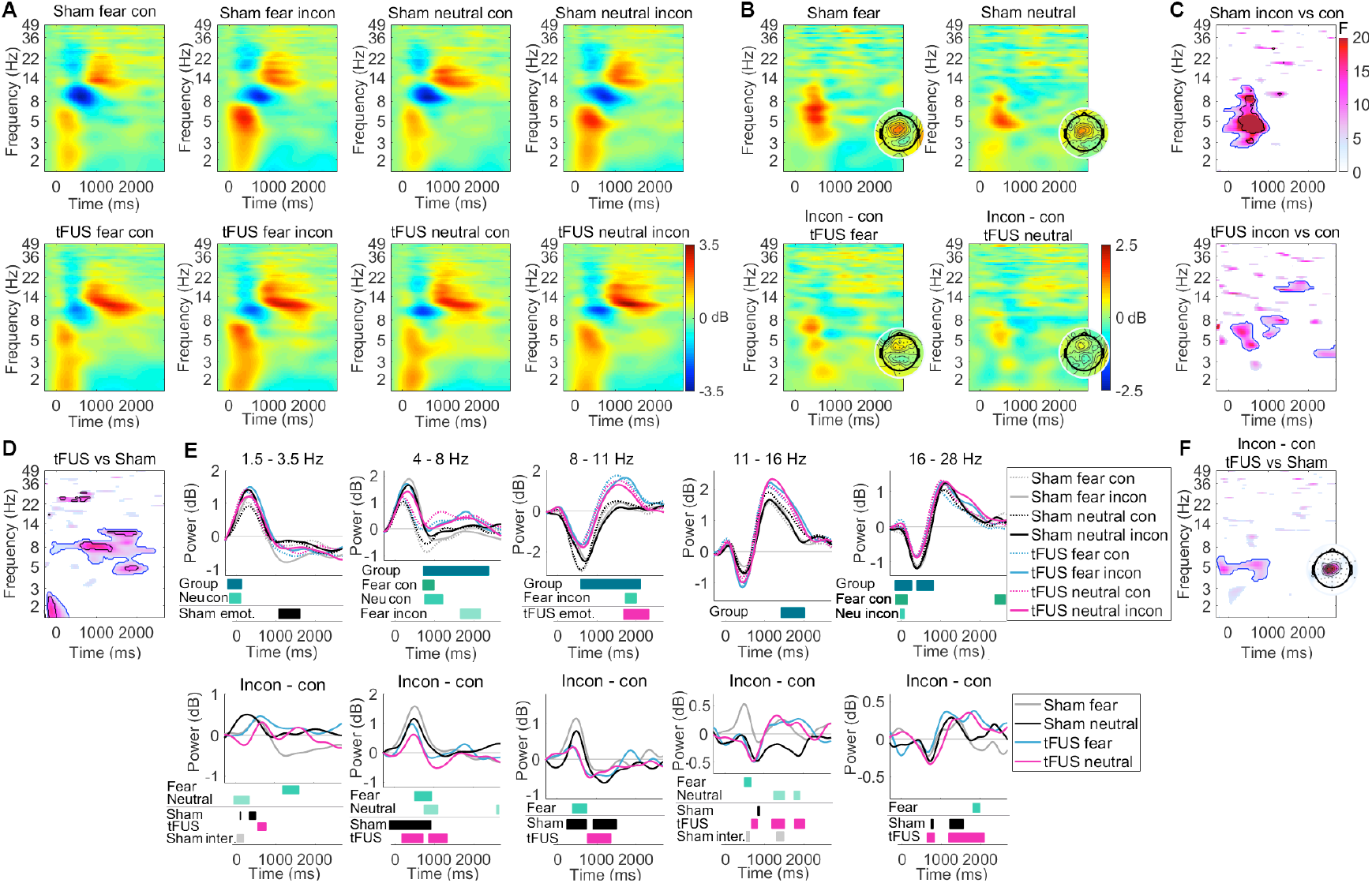
Frontocentral time frequency response with congruency contrasts. (**A**) ERSP at FCz for each condition (dB power change over baseline). (**B**) Incon – con contrast power for fear and neutral trials for each group. Inset displays scalp map of power (4 - 8 Hz; 200 - 800ms). (**C**) Significant main effect of congruency for each for each group (F - values). In **C, D**, and **F**, only significant values shows, blue outlines threshold-based clustering; black lines represent FDR correction (p < 0.05). (**D**) Significant main effect of group (RM-ANOVA). (**E**) Power in various frequency bands (top row); bars below represent significant differences across groups (Group: RM-ANOVA), permutation statistics displayed for each condition (green), and the main effect of emotion within groups (black and magenta). Incon – con contrast power is plotted below for each frequency band. Bars indicate significance across groups for each emotion (green). Additionally, the main effect of congruency for each group (Sham, tFUS), as well as any significant congruency × emotion interaction effects (inter.) are plotted (shades of black and magenta). (**F**) Statistical differences across groups (RM-ANOVA) for incon – con contrast power. Scalp map of F-value peak plotted in inset to the right (4-8 Hz; 0 – 800ms).

In the theta range, a strong and robust congruency effect was seen in the Sham group consistent with the literature on frontocentral theta and conflict monitoring tasks, however this effect was more diffuse and of much smaller amplitude in the tFUS group (Figure 4B and C). There was significant main effect of group in incon-con contrast power in the low theta range, most pronounced at central electrodes. Looking at frequency response over time (Figure 4E) demonstrates that incon-con contrast theta power was significantly smaller and of shorter duration in the tFUS group compared with sham, especially on fear trials. This is due to the fact that the Sham group showed a post-peak theta suppression while the tFUS group did not. Indeed in this time range, there were significant differences in theta power across groups in congruent trials for both face conditions.

In the alpha range, the tFUS group showed significantly less event locked alpha suppression than the Sham group, recovering faster to baseline and exhibiting a post suppression alpha activation, highest in the fear condition. Permutation testing showed a significant main effect of emotion in this time range (∼1800 ms). This alpha suppression was related to congruency processing as Sham group showed a congruency effect (earlier and larger peak suppression in congruency compared with incongruent trials producing a positive incon-con power peak). In contrast, the tFUS group showed smaller and faster suppression dynamics, so this effect was blunted; indeed there was a significant difference across groups in the incon – con alpha power in the fear condition.

In the low-beta range (11-16 Hz), there was no difference across groups in target-locked beta power suppression. However the Sham group did show reduced beta suppression in fear incongruent trails than all other trials, and a significant currency × emotion interaction at the low-beta suppression peak. Additionally, incon – con power was significantly larger in Sham than tFUS groups on fear trials. Furthermore, the post-suppression low-beta activation was larger and longer in the tFUS group. Liking higher in the beta range (16-28 Hz), there was significantly less target-locked power suppression in tFUS compared with Sham. Both groups showed significant congruency effects during the post-suppression beta recovery and subsequent peak. This peak was significantly longer in incongruent than congruent trials in the tFUS group, and significantly larger incon – con contrast power was observed in the tFUS group compared with Sham in the fear condition.

### Effects of dACC tFUS on frontal time-frequency response to fearful face distractors

Both the Sham and tFUS groups showed significant main effect of emotion (RM-ANOVA) in the low theta range (3.8 – 5.5 Hz) at frontal electrodes (Figure 5), however this effect begins earlier in the tFUS groups. In the 200 – 950 ms time range, the tFUS group showed a significant main effect of emotion, but the Sham group did not (Figure 5A). Comparing fear – neutral contrast power across groups, there was a significant difference between groups at frontal and parietal electrodes. However, at a later time window (1000 – 1900 ms), both groups showed a significant effect of emotion with no significant differences across groups. During the stimulus-induced low-theta peak, the tFUS groups showed a lower peak for fear than neutral in both congruency conditions, which was not the case for the Sham group, which showed no difference in congruent trials, and a slightly higher theta peak on fear than neutral trials in the incongruent condition (Figure 5B). Indeed, in this time range there was a significant main effect of group in fear – neutral contrast power, and a significant difference across groups with permutation testing in the incongruent condition. Additionally, a significant main effect of group was seen in the time range of 900 – 2500 ms. Following the initial theta activation, the tFUS group showed continued elevated power, greater in neutral than fear, and especially high in neutral congruent trials. Conversely, in the Sham group power returned to baseline with only a small elevation in power in the neutral condition. Significant differences were seen across groups with permutation statics in neutral congruent trials, and there was a significant congruency × emotion interaction effect in the tFUS group.

**Figure 5.**
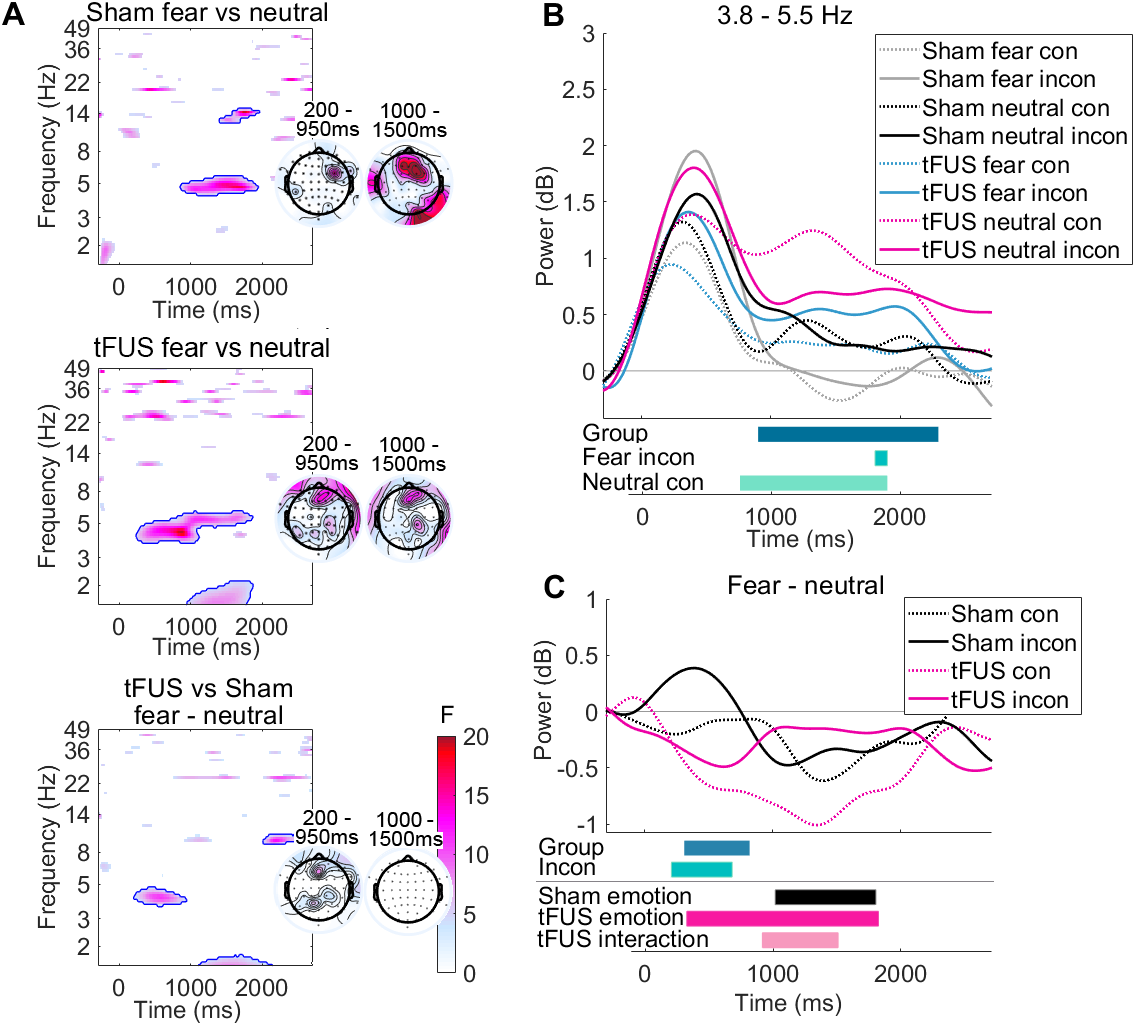
Groups differ in frontal theta during emotional face processing. (**A**) Main effect of emotion (fear vs. neutral, RM-AVOA) for Sham (top), tFUS (middle), and the main effect across groups for fear – neutral contrast power. Scalp maps in inset to the right of each plot represent significant differences across groups for power in the theta band (3.8 – 5.5 Hz) at 200 – 950 ms (left) and 1000 - 1900ms (right). (**B**) Event locked power in the low-theta band (3.8 – 5.5 Hz). The main effect of group (RM-ANOVA) and permutation statistics across each group for each trial condition are displayed below plot. (**C**) Fear – neutral contrast power. The main effect of group (RM-ANOVA) for fear – neutral contrast power (Group) and permutation statistics across groups for incongruent (incon) are displayed below. Additionally, the significant main effect of emotion and congruency × emotion interaction for theta power in B are displayed at the bottom of **C** (RM-ANOVA).

### Effects of dACC tFUS on event-related error response potentials

Baseline-subtracted ERPs were time locked to each subjects’ response (button-press) to assess response-locked potentials for correct and error responses. Both groups showed no difference across correct and error responses at the pre-response NPe peak, but showed a large frontocentral error-related negativity (ERN) and subsequent frontocentral positivity (Pe, ∼200 ms post response) for erroneous responses (Figure 6A and B). There were no differences across groups at NPe or peak ERN, but the ERN negative potential began earlier in the Sham than the tFUS group. Pe peak was significant larger amplitude and longer duration the Sham than tFUS group. Additionally, there was a significant group difference around 600 ms post-error where a fontal negativity and posterior positivity can be seen in the Sham group but not the tFUS group. Subtracting correct from error potentials produced an error – correct difference wave (Figure 6C and D). Again, no differences were seen at peak ERN, but onset was later in the tFUS group. Further, a larger Pe was seen in the Sham group (peak group differences in central and occipital electrodes). Finally, around 600ms there was a group difference in frontal and parietal-occipital electrodes.

**Figure 6.**
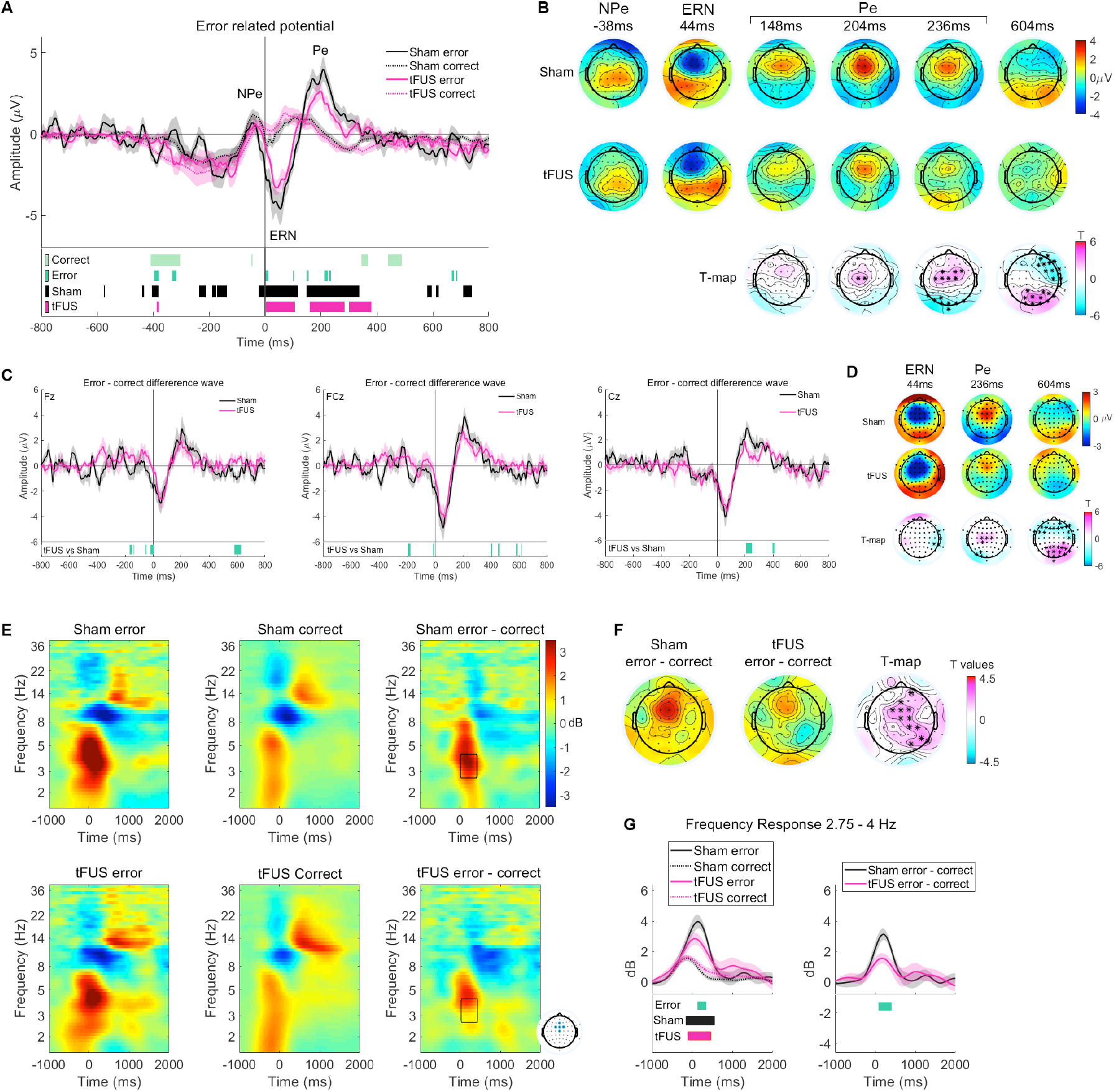
Transcranial FUS targeted to the dACC modulates event-related error response potentials. (**A**) Response-locked error-related potentials at FCz to correct and incorrect responses. Statistical significance across groups for correct and error trials, as well as error vs. correct for group (Sham, tFUS) (permutation testing) are displayed below plot. (**B**) Scalp maps correspond to peaks in A for Sham (top) and tFUS (middle). T values displayed at the bottom of the figure; asterisk (*) represents significant electrodes. (**C**) Error – correct difference potential (linear subtraction of correct from error response) for electrodes Fz, FCz, and Cz. (**D**) Scalp maps correspond to peaks in C and displayed as in B. (**E**) Error-related time frequency data. ERSP data for erroneous and correct responses, time-locked to the button press (0 ms). Data averaged across electrodes Fz, FCz, Cz, FC1, and FC2. Linear subtractions of correct from error power are also displayed (error – correct). The black boxes highlight region of statistical significance across groups, shown in further detail in **F** and **G**; the time (32 – 400 ms) and frequency range (2.75 – 4 Hz). (**F**) Error – correct contrast power for regions highlighted in **E** for each group. Scale same as E. Right-most map represents T values from permutation testing across groups as in **B**. (**G**) Frequency response over time in the delta band. Significant differences shown below are results of permutation testing across groups for error responses (Error), and within each group for error vs. correct responses (Sham, tFUS).

### Effects of dACC tFUS on error responses in time-frequency data

Response-locked ERSP data was calculated for both correct and error responses and pooled at frontocentral electrodes (Figure 6E). Both tFUS and the Sham group showed a larger delta and theta response on error compared with correct trials. Comparing error – correct contrast power across groups showed in the delta range this was significantly reduced in the tFUS group. Comparing the time and frequency range highlighted in figure 6E (2.75 – 4 Hz, 32 – 400 ms) a significant difference was seen across groups in both error (p < 0.001), and error-correct power (p = 0.009). Scalp maps indicate that the topography of the significant differences is both frontocentral and right parietal. Additionally, the tFUS groups show less post-error alpha suppression on correct than error trials.

### Influence of dACC tFUS on heart rate and heart rate variability

In order to assess physiologic changes in response to emotion face distractors and tFUS stimulation, heart rate was measured and several heart rate variability (HRV) metrics were calculated: standard deviation of normal-to-normal heartbeat (SDNN), percent of successive normal R-R intervals exceeding 50 ms (pNN50), short term HRV (SD1, also known as the root mean square of successive R-R interval differences, RMSSD), long term HRV (SD2), and SD1/SD2 ratio (see Methods). Three-minute time windows were selected for analysis: the first taken towards the end of the baseline trials (while subjects were performing a simple flanker, no faces, no stimulation), and the second being the first three minutes of the main trials (fearful and neutral faces appeared behind flanker distractor arrows, and stimulation group received online tFUS to the dACC, Figure 7). Mixed-measures, two-way RM-ANOVA’s [2 groups × 2 time points] show a group × time interaction effect for HR [F_HR_(1,24) = 7.74, p = 0.010, η_p_^2^ = 0.24], as well as all HRV measures: SDNN [F_SDNN_(1,24) = 54.23, p < 0.001, η_p_^2^ = 0.69], pNN50 [F_HR_(1,24) = 9.72, p = 0.005,, η_p_^2^ = 0.29], SD1 [F_SD1_(1,24) = 5.80, p = 0.024, η_p_^2^ = 0.19], SD2 [F_SD2_(1,24) = 51.7, p = < 0.001, η_p_^2^ = 0.68], and SD1/SD2 ratio [F_SD1/SD2_(1,24) = 4.31, p = 0.049, η_p_^2^ = 0.15] (Table S4).

**Figure 7.**
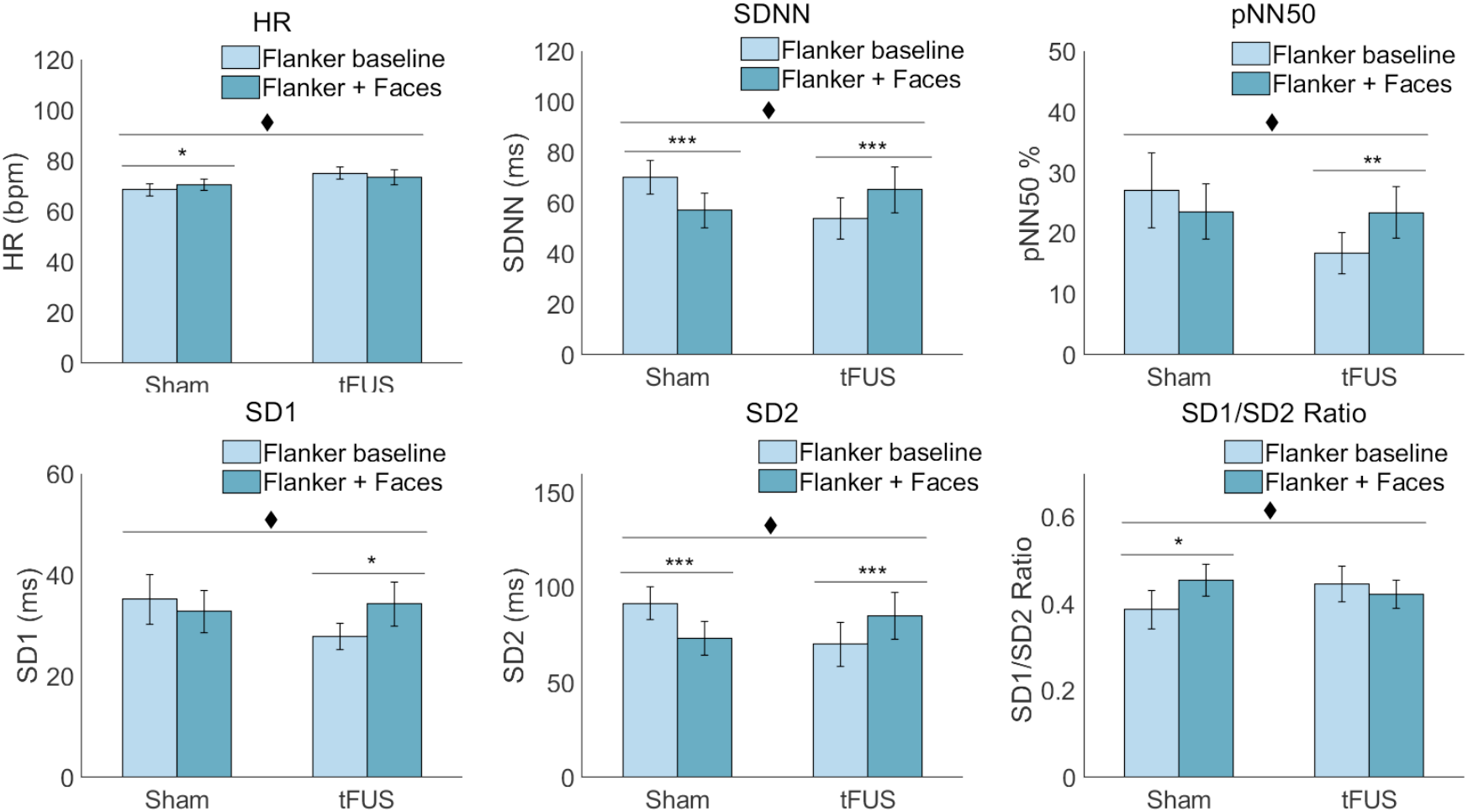
Heart rate and heart rate variability changes with addition of emotional face distractors. HR metrics were calculated from three minute samples recorded at two different time points: baseline (flanker only, no stimulation), and during the first three minutes of the main trials (onset of emotional faces and tFUS). The significant group × time interaction effect (♦p < 0.05), and post-hoc statics are displayed. All post-hoc statistics are Bonferroni-corrected (* p < 0.05, **p < 0.01, ***p < 0.001). Related to Table S4.

Post-hoc analysis shows at the onset of emotional faces the Sham group significantly increased heart rate (↑3 ± 1%, p = 0.037, mean ± SEM), while there was a small but not significant decrease in HR seen in the tFUS group (↓2 ± 1%, p = 0.098). Additionally, all measures of HRV decreased in Sham group at the onset of faces with significant decreases in SDNN (↓20 ± 4%, p < 0.001) and SD2 (↓22 ± 4%, < 0.001). Non-significant decreases were seen in pNN50 (↓9 ± 11%, p = 0.150) and SD1 (↓4 ± 6%, p = 0.37). Conversely, HRV significantly increased in the group receiving tFUS to the dACC measured in the SDNN (↑24 ± 5%, p < 0.001), pNN50 (↑64 ± 18%, p = 0.007), SD2 (↑25 ± 6%, < 0.001), and non-significantly in SD1 (↑21 ± 7%, p = 0.37). The Sham group significantly increased in SD1/SD2 ratio (↑25 ± 9%, 0.039), while the tFUS group showed no change (0 ± 8%, p = 0.51). There were no significant main effects of time (F < 2.91, p > 0.10) or group (F < 1.86, p > 0.19. Unpaired t-test shows no significant differences between groups at baseline (all T ≤ 1.96, p ≥ 0.062).

### Effects of transient tFUS sessions on acute mood

Subjects completed the PANAS mood questionnaire at baseline and immediately following experiment completion. Ratings were summed across positive and negatively valence probes to create PANAS positive and negative scores (Table 1). Score were assessed across group and time using two-way mixed measures RM-ANOVA [2 groups × 2 time points (baseline, post-experiment)]. PANAS negative scores showed no significant main effect of time, group or interaction effect (F < 2.52, p > 0.12). For PANAS positive scores however, there was a significant main effect of time [F_Positive_(1,26) = 13.99, p = 0.001, η_p_^2^ = 0.35] indicating positive PANAS scores decreased post experiment (baseline: 28.9 ± 14, post: 24.2 ±1.7, mean ± SEM). There was no main effect of group [F_Positive_(1,26) = 2.49, p = 0.19, η_p_^2^ = 0.09] or group × time interaction [F_Positive_(1,26) = 0.85, p = 0.37, η_p_^2^ = 0.03]. Post-hoc tests revealed that although both groups showed decreased PANAS scores, only the Sham group decreased significantly (Sham: p = 0.003, tFUS p = 0.057). Both groups showed a decreased score for ‘interested’ (Sham p < 0.001, tFUS p = 0.002), but only the Sham group showed significant decreases in scores for ‘excited’, ‘enthusiastic’, and ‘attentive’ (p = 0.003, 0.002, 0.007; tFUS p’s > 0.21).

**Table 1.** Summary of PANAS mood data. PANAS data collected immediately prior to (Baseline) and following completion of experimental session (Post). Statics represent post-hoc paired t-tests performed within groups. All p-values Bonferroni-corrected. All values displayed as mean ± SEM (*p < 0.05).

|  | Sham |  |  | tFUS |  |  |
| --- | --- | --- | --- | --- | --- | --- |
|  | Baseline | Post | Baseline vs. post | Baseline | Post | Baseline vs. post |
| PANAS positive | 31.6 ± 1.9 | 25.9 ± 2.7 | 0.003* | 26.1 ± 1.9 | 22.6 ± 2.1 | 0.057 |
| PANAS negative | 13.8 ± 1.1 | 13.5 ± 1.1 | 0.64 | 11.5 ± 0.4 | 12.1 ± 0.6 | 0.31 |

### Effects of dACC tFUS on reaction times

To rate task performance, a Mann-Whitney U test was performed across groups on baseline-subtracted median RTs (baseline congruent RT was used for congruent trials, and likewise for incongruent trials) (Figure 8). The results indicate that in both the neutral and fear incongruent condition, baseline-subtracted RT was faster in the tFUS group (Mdn = −3 ms, −3ms) than the Sham group (Mdn = 13 ms, 7 ms), (fear: U = 48.5, p = 0.021, neutral: U= 54.5, p = 0.044). No Significant difference was found in the neutral, fear, or oddball congruent condition (U ≥ 76.0, p ≥ 0.39) or oddball and post incongruent condition (U ≥ 74.0, p ≥ 0.285). Furthermore, more subjects demonstrated faster RTs than baseline in the tFUS group in all conditions except the post congruent (Supplementary Table S5).

**Figure 8.**
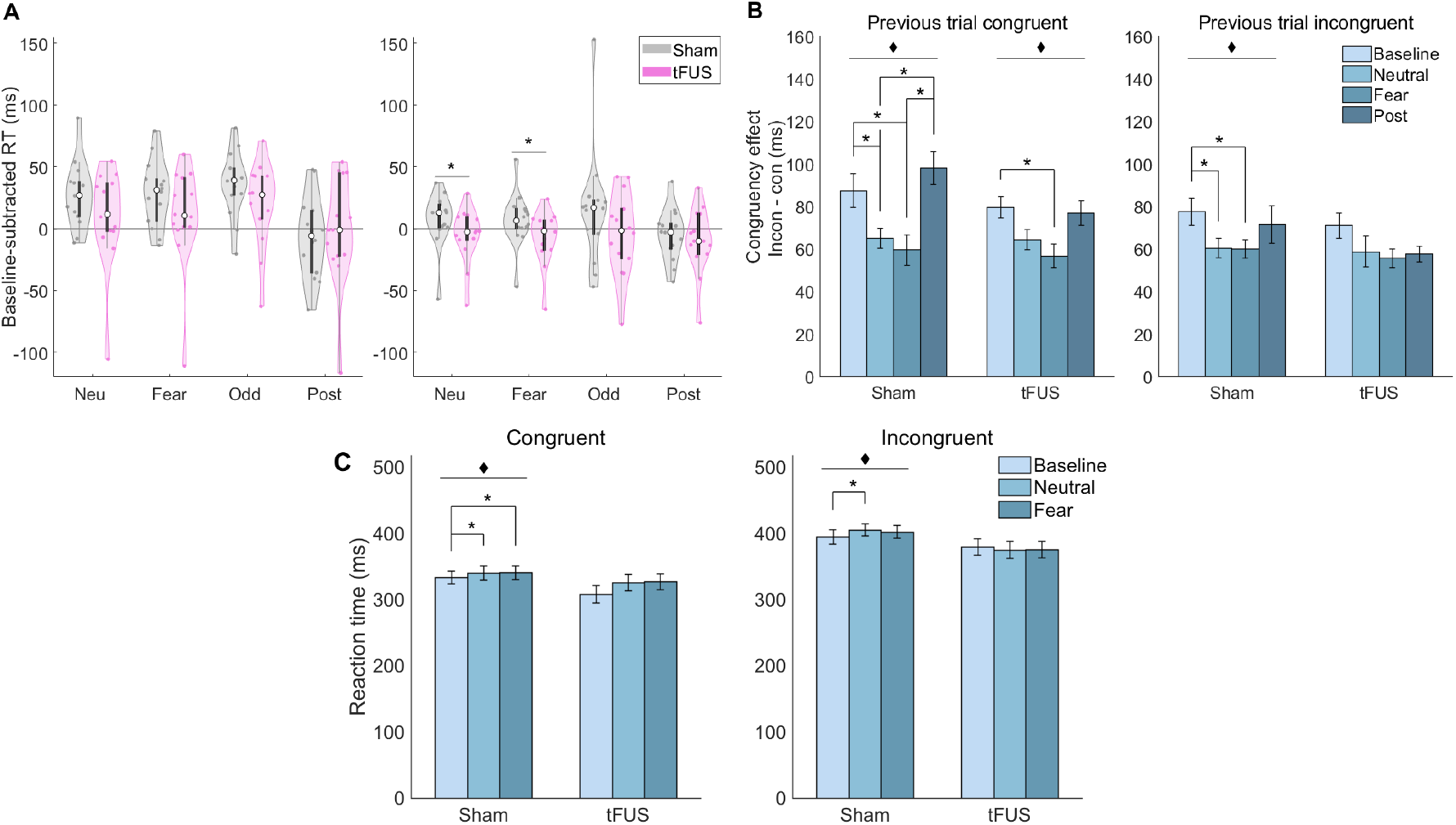
Reaction Time and congruency effect. (**A**) Baseline-subtracted reaction times displayed for both congruent (left) and incongruent (right) trials. Box-plots overlaid on violin plots (white circle at median), statistics represent the results of Mann-Whitney tests across groups *p < 0.05. (**B**) Congruency effect for RT separated by previous trial congruency. Statics represent the results of related-samples Friedman’s analysis (♦p < 0.05) and post-hoc tests (*p < 0.05, Bonferroni corrected for multiple comparisons). (**C**) Uncorrected reaction times compared within group with nonparametric statics as in **B**.

Using non-parametric Friedman’s test to compare median RTs within groups further supports this finding (Figure 8C). To assess the behavioral effect of emotional face distractors on reaction time, RTs in baseline, neutral and fear trials were compared within group for each congruency condition. In congruent trials, the RT in the Sham group significantly differed across baseline, neutral, and fear RT (χ^2^(3) = 9.93, p = 0.007), but the tFUS group does not (χ^2^(3) = 4.86, p = 0.089). Post-hoc tests show that in the Sham group both congruent fear (p = 0.014) and neutral (p = 0.032) were significantly slower than baseline. Similarly, in the incongruent condition the Sham (χ^2^(3) = 7.00, p = 0.030), but not the tFUS group (χ^2^(3) = 1.78, p = 0.41) showed a significant slowing of RTs with the addition of the faces. Post-hoc tests show that after Bonferroni correction, only the slowing in the neural condition was significant in the Sham group (fear: p = 0.113, neutral: p = 0.042).

### Effects of dACC tFUS on conflict adaption

Additionally, to assess conflict adaption and its interaction with emotion, the congruency effect (also known as the flanker effect: incongruent RT – mean congruent RT) was calculated. RTs were separated by the congruency of the previous trial, and congruency effects were calculated and compared within groups (Figure 8B). On trials following congruent trials, a significant difference in congruency effect between baseline, fear, neutral, and post-stimulation trials is seen in the Sham group (χ^2^(3) = 19.58, p < 0.001), and tFUS group (χ^2^(3) = 11.14, p = 0.011). Bonferroni-corrected post-hoc statistics show that in the Sham group neutral trials have a significantly smaller congruency effect than baseline (p = 0.016) and post-experiment (p = 0.002), while in fear congruency effect is significantly smaller than post experiment (p = 0.010), and substantially smaller than baseline (p = 0.062). In the tFUS group there only a significantly smaller congruency effect on fear compared with baseline (p = 0.05).

On trials following incongruent trials however, the Sham group has a significantly smaller congruency effect for trials with distractor faces (χ^2^(3) = 13.10, p = 0.004) while this is not the case for the group that received tFUS to the dACC (χ^2^(3) = 3.62, p = 0.35). Post-hoc tests show Sham congruency effect is significantly larger at baseline than on trials with fear (p = 0.016) or neutral face distractors (p = 0.041).

### Effects of dACC tFUS on post error slowing and accuracy

To assess post error slowing, RTs on fear and neutral trials following an error were compared with median RTs in using Wilcox signed rank tests. The Sham group showed significant post error slowing on fear trails in both flanker conditions (congruent: Z= 77, p = 0.028, incongruent: Z= 78, p = 0.023). The tFUS group showed post error slowing in both incongruent conditions (fear: Z= 66, p = 0.034, neutral: Z=77, p = 0.028). All other conditions were not significantly slower than baseline Z < 71, p > 0.24).

To assess post error slowing across groups, median RT was subtracted from post error RTs. There were no differences across groups in post error RT (U ≥ 27, p > 0.2). Additionally, there were no differences in either group in post error slowing based on current trial distractor face emotions (Z < 66, p > 0.15). Mann-Whitney U tests revealed no significant differences across groups in response accuracy neither at baseline, nor during the experiment (U ≥ 68.0, p ≥ 0.178, Table S6).

## DISCUSSION

The dACC is highly involved in numerous tasks involved in directing executive control. Previous research has implicated it in attention, cognitive control, conflict, error, reward, interoceptive awareness, emotion, pain, and relaxation (Critchley et al., 2002; Critchley et al., 2004; Shackman et al., 2011; Vogt, 2005). Both functional imaging and electrophysiological data have linked activity in the dACC with performance on cognitive attention tasks (Matthews et al., 2007; Weissman et al., 2006), as well as emotional interference (Shafer et al., 2012). Furthermore it is involved in concentration meditation, as well as emotional awareness (McRae et al., 2008). Indeed, attentional lapses (as measured by negative performance on cognitive interference tasks) are correlated with reduced pre-stimulus and evoked activity in the cingulo-opercular network (Kerns et al., 2004; Weissman et al., 2006).

Given the central role of the dACC in cognitive control, the aim of this study was to examine the feasibility of modulating the dACC and broader DMN activity using MR-targeted tFUS. Our observations provide robust evidence that MR-targeted tFUS produced significant changes in neurophysiological and neurobehavioral conflict processing, emotional attention, and cognitive control consistent with dACC and DMN modulation.

### Elimination of reaction time slowing in response to emotional face distractors and alteration of conflict adaption by dACC tFUS

As was expected based on previous findings (Jasinska et al., 2012; Papazacharias et al., 2015), the group that received Sham showed a significant slowing of RTs from baseline with the addition of neutral and fearful faces as distractors behind the flanker task (Figure 1), yet no significant difference was seen in the tFUS groups (Figure 8C). Additionally, comparing baseline-subtracted RTs across groups, the tFUS group was significantly faster in fear and neutral incongruent conditions (Figure 8A). Response slowing induced by emotional faces is known to recruit the cingulo-opercular network (Papazacharias et al., 2015), and these results suggest that this was disrupted by tFUS to the dACC, resulting in a reduced distraction impairment of RT performance.

To study conflict adaption, congruency effect (incongruent-congruent RT) was separated by previous trial congruency and compared within groups. Consistent with previous studies, the Sham group showed significantly reduced congruency effect for trials with emotional distraction in both trials following congruent and incongruent trials (Egner et al., 2007). However, the tFUS group only differed from baseline on trials following congruent trials and no difference from baseline on trials following incongruent trials. Previous studies have shown that congruency effect is reduced in negative mood (van Steenbergen et al., 2010). This result suggests that tFUS to the dACC may bias conflict adaption, modulating conflict processing by enhancing sensitization to conflict on trials following incongruent trials.

### Event-related potentials show enhanced early components and modulated fear processing by dACC tFUS

Individuals receiving tFUS to the dACC showed an earlier onset and larger amplitude early frontocentral negativity and parietal positivity following presentation of the distractor faces and arrows (D-N1, ∼ 100 ms). The N1 component is associated with the orienting network, is known to decrease after attention fatigue (Boksem et al., 2005). At D-P1 (the first frontocentral positivity), the sham group shows a greater amplitude peak for fear than neutral distractor trials consistent with the literature (Carlson and Reinke, 2010), yet this is not the case for the tFUS groups, and the two groups differ significantly in fear – neutral potential at frontocentral electrodes. This ERP peak is thought to facilitate spatial attention through involvement of the amygdala, dACC and visual cortex (Carlson et al., 2009; Klumpp et al., 2012), and is sensitive to fear arousal (Dennis and Chen, 2007). Additionally, the Sham group showed no difference across conditions at the following frontocentral negative peak (T-N1), but the tFUS group shows a significantly pronounced negative potential for fear compared with neutral distractors. It is possible that tFUS to the dACC modulated this emotional appraisal signal, which perhaps had broader implications to reduced distraction effects described above in RT.

There were no significant differences in N2 peak amplitude across groups (Figure 2), yet comparing incon – con contrast is was clear that the tFUS group had an earlier onset of N2 than Sham on fear trials (this has also been observed in mindfulness meditators (Fan et al., 2015)). Additionally, the tFUS group had a diminished incon – con response at P3 compared with Sham. Previous research has shown P3 in distractor processing is higher in novices compared to meditators (Cahn and Polich, 2009) and was reduced with mindfulness meditation training (Moore et al., 2012), and that faster RTs in meditators on incongruent trials can be correlated with changes in P3 (Jo et al., 2016). Frontal P3 is presumed to come from the dACC, is evoked by attention-switching (Xie et al.), and is emotion dependent (Albert et al., 2010). This suggests that tFUS perhaps enhanced sustained attention and reduced the need for attention switching.

### Modulation of time-frequency EEG data across multiple frequency by dACC tFUS

The tFUS group had an earlier onset of stimulus-induced frontocentral delta, and a longer sustained delta activation at parietal electrodes than Sham. Both groups show differing congruency × emotion interactions in the delta band at parietal electrodes. In the theta band, there were no differences across groups in target-induced theta peak activation, however the Sham group displayed a post-peak theta suppression, while the tFUS group showed a second theta which was highly significant over the sham group in parietal-occipital electrodes and to a lesser extend frontal electrodes. Additionally, differences were seen across groups in the incon – con contrast power, which was reduced in tFUS subjects in theta band due to sustained activation of theta. Frontocentral theta is known to be present both in cognitive tasks and meditation (Inanaga, 1998), and is thought to originate from the dACC and functionally connects to other regions of the brain for executive control of action updating (Cohen, 2011). It is linked to error monitoring (Cavanagh et al., 2009), conflict adaption (Cohen and Cavanagh, 2011), and theta coherence is modulated by reaction time (Cavanagh et al., 2009). Central delta and frontocentral theta are both associated with N2 and P3 in response inhibition, but thought to indicate separate processes (Harper et al., 2014).

Additionally, subjects receiving tFUS to the dACC showed significantly reduced alpha suppression and subsequent post trial alpha activation at frontal and parietal electrodes. Differences were seen across groups in the incon – con contrast power, in the alpha band due to reduced target-locked alpha suppression on congruent trials in the tFUS group. Alpha is related to tonic alertness and suppression is involved in ignoring emotional distractors, increases with increasing distractor frequency (Murphy et al., 2020). Failure to suppress alpha in visual and sensorimotor areas predicts error and decreased performance in sustained attention (Mazaheri et al., 2009).

It has been proposed that the cingulo-opercular network involving the dACC and insula, maintain tonic alertness through alpha oscillations, alpha is negatively correlated with activity in the dorsal attention network and attention is allocated to this network when necessary by disrupting alpha oscillations (Sadaghiani et al., 2010). Additionally, elevated midfrontal theta and parietal alpha power are associated with increased awareness of conflict (Jiang et al., 2015). The data presented here suggests that tFUS to the dACC modulated task-related alpha suppression, as well and theta congruency processing which could explain the reduced reaction time slowing observed in the tFUS group.

### Modulation of error responses by dACC tFUS at Pe and in the delta EEG band

The tFUS and Sham did not differ in peak ERN amplitude, although the tFUS group did show a slightly delayed ERN onset latency. However, the following error-related frontocentral positivity (Pe), was smaller in amplitude and duration in the tFUS group. Pe represents different aspects of error processing from ERN and may reflect conscious recognition of an error (Endrass et al., 2007; Overbeek et al., 2005) with a motivational significance.

Significant differences were seen across groups in error responses in the delta EEG range (1.5 – 4 Hz). The tFUS group showed less delta power than the Sham group in frontocentral electrodes on errors as well as in error – correct contrast power, but no differences were seen across groups in the theta range. Delta and theta combined are related to the ERN (Munneke et al., 2015) but may represent separate processes. The delta band is primarily associated with performance monitoring and error detection, while theta band activity may be associated more with motor execution failure (Cohen and Cavanagh, 2011; Yordanova et al., 2004), and correlated with N2 (Cavanagh et al., 2017). Others have shown that response-locked delta-band phase coherence may support general cognitive function (Cavanagh et al., 2009), decision making, and saliency (Knyazev, 2007).

Additionally, the tFUS group shows significant reduction of alpha power post error compared with correct responses, related to adaption after errors (van Driel et al., 2012), but the Sham group does not. Combined fMRI EEG research and lesion studies suggests that the dACC may not be the generator of N2 but is likely is the generator of the error related ERN (Iannaccone et al., 2015; Stemmer et al., 2004). These findings suggest that MR-targeted tFUS delivered to the dACC modulated error processes perhaps through delta biasing, potentially affecting emotional awareness or cognitive control process related to error recognition.

### Heart rate increases and heart rate variability decreases associated with presentation of emotional face distractors are attenuated by dACC tFUS

A significant interaction group × time interaction effect was seen for physiologic response to fear and neutral face distractors in heart rate and HRV. The Sham group significantly increased heart rate and decreased SDNN, SD2, and SD1/SD2 ratio, while the dACC tFUS group substantially decreased heart rate, and significantly increased SDNN, pNN50, SD1, and SD2. Previous studies have shown that HRV decreases and HR increases with negative faces and increasing load (Park et al., 2014), exactly as was observed in the Sham group here.

The SDNN, influenced by sympathetic but largely parasympathetic activity, is known to decrease with increased workload (Fallahi et al., 2016) and increase with slow relaxed breathing (Shaffer et al., 2014). SD1 (also called the RMSSD) is associated with short-term HRV and vagal modulation of HRV (Shaffer et al., 2014), whereas SD2 is thought to measure both short and long term HRV and correlated with baroreflex sensitivity (Guzik et al., 2005). SD1/SD2 is associated with autonomic balance. Previous studies have shown that SD1 and SD2 increase with frontocentral theta power during concentration of awareness on the breath in Zen meditation (Kubota et al., 2001). The pNN50, like the SD1 is largely influenced by parasympathetic activity (Shaffer and Ginsberg, 2017).

A Meta-analysis of neuroimaging studies suggests that HRV can provide an index of top-down appraisal of threat, and that cortical substructures including the anterior cingulate influence the body’s autonomic response by connections with the insula, and that the perceptual experiences of threat and safety are linked to HRV via the cingulate (Thayer et al., 2012). The above-described results indicate that tFUS to the dACC both inhibited physiologic responses to fearful and neutral distractor faces, but perhaps enhanced states of relaxation and parasympathetic activation. Since lower HRV is associated with an impaired ability to inhibit attention from fearful faces (Park et al., 2012), it is possible that tFUS causally affected reaction times by modulating fear processing.

### Conclusions

The dACC is known involved in task switching and error (Nee et al., 2011), but its direct role in conflict processing is debated, and it is now thought not to be the direct generator of N2 (Iannaccone et al., 2015; Nee et al., 2011). This is consistent with the results presented here, in which tFUS targeted to the dACC did modulate conflict processing but failed to produce a direct effect on N2.

The dACC is known to be active in states of relaxed concentration and meditation (Hölzel et al., 2007; Kubota et al., 2001). Our observations demonstrate that MR-targeted tFUS delivered to the dACC produced a hallmark effect that might be expected from relaxed contention, including reduced RT performance reduction in response to fearful face distractors and an increase rather than a decrease in parasympathetic markers of the HRV. This suggests that tFUS altered emotional processing and enhanced sustained attention perhaps by reducing attentional engagement with emotional faces and thereby reducing the need for attention switching evidenced by early ERP components, P3, elevated post-trial theta, reduced alpha suppression, and modulation of delta. In summary, our observations provide evidence that tFUS targeted to a single brain area, such as the dACC, can be used for broader DMN modulation in a manner that has numerous, practical therapeutic applications in neuropsychiatry and mental health.

## MATERIALS AND METHODS

### Human Participants and Informed Consent

All procedures were performed using protocols approved by the Arizona State University Institutional Review Board. The study recruited 28 healthy, right-handed subjects from the university community (ages 19–39). Following screening, human volunteers were provided with a final overview of the study prior to providing informed consent. All participants reported no history of neurological or psychiatric disorders, no hearing or uncorrected visual impairments, migraines, or medication use, as well as no contraindications for MRI.

Subjects were randomly assigned to either the sham group (control) or the stimulation group (tFUS). The sham group (n = 14) included 3 women and 11 men, mean age 22.4. The tFUS group (n=14) included 5 women and 9 men (mean age 25.3). One subject in each group was removed from EEG and HR analysis due to poor quality data recording.

### Behavioral task protocol: modified flanker paradigm with emotional face distractors

Subjects performed a modified version of the Eriksen Flaker task (Eriksen and Eriksen, 1974). After the experimental setup and completion of the mood surveys, subjects began with 32 practice trials, and then performed 100 baseline trials (50 congruent, 50 incongruent). Practice and baseline trials consisted of a simple flanker task with white distractor arrows appearing on a black background for 100 ms, followed by the target arrow. The distractor arrows were either congruent (pointing in the same direction as the target) or incongruent (pointing in opposite direction of the target). Subjects were asked to report the direction of the target arrow via button press with the index (left) or middle (right) finger of their right hand; participants were instructed to respond as quickly as possible without sacrificing accuracy. All main experimental trials utilized the protocol displayed in Figure 1. Each trial began with a fixation cross which was presented for 1800-2500 ms. Both groups were presented with a sham sound meant to imitate the sound of the tFUS pulse that is herd through bone conduction. This sound began in both groups 28 ms prior to the appearance of the distractor arrows on the screen, and the tFUS stimulation (described below) began at the same time. Both the sound and the stimulation had a duration of 500 ms. Distractor arrows then appeared overlaid on one of three face types: a face exhibiting a neutral expression, one exhibiting a fearful expression, or a scrambled face (colored static). The fearful and neutral faces appeared with equal frequency, while the scrambled trials occurred only 3/50 as an oddball. Trials were presented in four blocks of 105 trials. Trials were pseudo-randomized with equal number of each trial type, and arrow direction. Face images were used from both the NimStim (Tottenham et al., 2009) and MaxPlank (Ebner et al., 2010) (young faces only) datasets. Faces images were edited to have a black background, masked to include just the face, and resized where necessary to ensure consistency in face size as well as position of nose and eyes.

### Experimental setup

The task, tFUS stimulation, and all audio and visual information were controlled using PsychoPy software (Peirce, 2007). To ensure accurate notation of timing distractor and target presentation in the EEG file, events were triggered through photodiodes on the presentation computer monitor using Cedrus StimTracker (Cedrus Corporation, San Jose, CA). Likewise, all responses were recorded using a response box. Sham sound and tFUS stimulation were controlled also by the presentation computer via parallel port, and split to record triggers in the EEG system.

### Sham tFUS Procedures

In both the sham and the stimulation group the ultrasound transducer was placed on the head and all ultrasound equipment turned on. For subjects receiving sham the tFUS transducer was disconnected from the RF amplifier. Both groups wore headphones, and at the onset of the tFUS stimulus, a low-volume, high-pitched sound was played to simulate the sound of the tFUS pulses that is herd through bone conduction. The sound file was created by combing a very high frequency tone with a square wave at the pulse repetition frequency. Upon questioning subjects in the tFUS group following the experiment, not perceptual difference was noticed between the sham sound and the tFUS preserved PRF pulse.

### MRI acquisition and processing

Structural T1 MRI scans of each subject in the tFUS group were collected prior to experimentation for the purposes of neuronavigation. Images were collected in a Philips Ingenia 3T scanner with a 32-channel head coil, using a 3D MP-RAGE sequence (TR = 2300 ms, TE = 4.5 ms, 1 x 1 x 1.1 mm^3^ voxels, field of view 240 x 256 mm^2^, 180 sagittal slices). Brainsight neuronavigation system (Rogue industries) was used to plan stimulation targets and guide placement of the transducer beam profile with respect to each individual’s anatomy. Montreal Neurologic Institute (MNI) coordinate system (Evans et al., 1994) was warped to each subject’s brain, and when planning the tFUS target both MNI coordinates and individual anatomy of the dACC was considered. A mean stimulation location of x = 2.9 ± 0.8, y = 22.2 ± 1.7, z = 32.8 ± 1.6 (mean ± SEM) was recorded (Figure 1).

### tFUS stimulation

Participants were blinded to mode of stimulation they received (active vs. sham). Following EEG setup, an infrared optical tracking system (Polars Vicra, NDI Medical, Waterloo, Ontario, Canada) was used to register the subjects’ structural MRI scans in virtual space, with their head and the ultrasound transducer in 3D real space. This allowed the transducer to be positioned correctly on the surface of the scalp in order to hit the anatomical target identified prior. A custom-built 3D printed housing was made for the transducer to hold the optical tracking unit and silicon spacer (ss-6060, Silicon Solutions, Cuyahoga Falls, OH), which was used to achieve the desired focal depth and couple the transducer to the scalp. Acoustic conductive gel was applied to both the transducer and the scalp. After correct placement of the transducer using the neuronavigation, we recorded the coordinate of the stimulation target. The transducer was held flush to the head with a custom-made, lightweight, elastic mesh cap, which did not interfere with EEG recording.

A broadband, single-focus transducer with a lateral spatial resolution of 4.9 mm^2^ and axial spatial resolution of 18 mm^2^ was used for this study (Blatek, Inc., State College, PA, USA). Given that tFUS is capable of inducing event related activity (Dallapiazza et al., 2017; Lee et al., 2016a; Lee et al., 2016b), all ultrasound stimulation was be delivered online, in a trial-by-trial manner. Each trial, stimulation began at the onset of the distractor arrows and face image. Each tFUS pulse had a carrier frequency of 0.5 MHz (to optimize signal transmission across the skull (Tufail et al., 2010), a pulse repetition frequency (PRF) of 1000 Hz, a 24% duty cycle, and duration of 500ms. Stimulation was triggered by the experiment PC, controlled through a two-channel, 2 MHz function generator (BK Precision, Edison, NJ) and driven by a 40 W linear RF amplifier (E&I 240L; Electronics and Innovation, Rochester, NY, USA) as described preciously (Legon et al., 2012). Water tank measurements indicated a max pressure of 1.0 MPa and spatial peak pulse average intensity (I_sppa_) of 20.4 W/cm^2^ at the focus.

### EEG recording

Electroencephalography (EEG) data was recorded using a 64-channel ActiCap system (BrainVision, Morrisville, NC, USA) with the standard 10-20 electrode layout. Electrode AFz was removed for placement of the ultrasound transducer. Data was recorded using a sampling rate of 5 kHz, resolution of 0.1 V, and band-pass filter of 0.1-100 Hz. The ground was placed at FPz and reference at the left mastoid. EEG electrode locations were recorded with Captrack camera system (BrainVision); fiducials were placed on the left and right tragus, and nasion.

### EEG Processing

EEG data was processed using custom scripts in MATLAB R2017b (MathWorks, Natick, MA, USA) with the utilization of EEGLAB v14.1.1 (Delorme and Makeig, 2004). Raw data was first down-sampled to 250 Hz and high-pass filtered at 1 Hz, and notch filtered at 60Hz. Data was then visually inspected for artifacts and bad channels were removed. Additionally, any channels with absolute temporal standard deviation greater than five or that exhibited artifacts for greater than 25% of the recording session were removed. All removed channels were then interpolated and all data was re-referenced to the scalp average using individually-recorded electrode locations. Independent Components Analysis was used to remove eye movement, blink, and other glaring artifacts; on average 3.6 ± 0.4 components were removed per subject.

### Event related potentials

Data was then epoched around the distractor arrow presentation (target-locked) and 200 ms pre-stimulus baseline was subtracted to create event related potential (ERP) data. A linear subtraction of congruent from incongruent ERP data was performed to assess the incongruent-congruent (incon – con) difference potential. For analysis of response-locked data, ERP data was then time-locked to the button press (response). In order to control for multiple comparisons problems, ERP data was analyzed using nonparametric permutation statistics when comparing across groups and within groups for comparing only two conditions. Statistical p values represent the proportion of 1,000 permutations of randomly shuffled data which produce a t value greater than that calculated by a standard two-tailed t-test (Maris and Oostenveld, 2007) (p < 0.05 was considered statically significant). Two-way RM-ANOVA [2 congruency conditions (congruent, incongruent) × 2 face emotions (neutral, fear)] were used to compare within groups.

Additionally for each individual, peak-to-peak amplitude and latency were identified for each the ERP complex, and compared across groups (Table S2 and S4). Additionally nonparametric Friedman’s test was used to compare within groups (Table S3).

### Error-related potentials

To analyze response-locked ERPs for correct and error responses, baseline-subtracted data was then time-locked to the response (button press). As with target-locked ERPs, each condition was compared across groups using permutation testing. Additionally, error and correct responses were compared within group using permutation testing (paired rather than unpaired t-tests were used).

### Time – frequency analysis: Event-related spectral perturbation

Event-related spectral perturbation (ERSP) data for time-frequency analysis was computed on time-locked data using EEGLAB. A 500 ms pre-stimulus baseline was used. Morlet wavelets were used with 3 cycles at the lowest frequency (1.5 Hz), and increasing linearly to 40 cycles at the highest frequency (50 Hz). All results are displayed as decibel power above baseline.

Similar to ERPs, permutation testing was used on each time and frequency data point. ERSPs were further quantified across groups with mixed measures RM-ANOVAs [2 groups × 4 trial conditions (fear congruent, fear incongruent, neutral congruent, neutral incongruent)]. On main trials within group analysis was done using RM-ANOVAs [2 congruency conditions (congruent, incongruent) × 2 face emotions (neutral, fear)]. For comparing error with correct responses, permutation statistics were used for across (unpaired) and within (paired) group analysis.

A cluster-based multiple comparisons correction was applied to statistical results by determining clusters of contiguous significant pixels, F or t values in these clusters were then summed and only clusters greater than 2 standard deviations above the mean were retained. Additionally, false discovery rate (FDR) correction was also applied. Specific frequency band and time windows were selected for analysis as scalp maps and frequency response over time based either on peak responses in the ERSP or the results of group level statistical testing.

For main trails, primary analysis was done only on neutral and fear, congruent and incongruent trials. Linear subtraction of correct from error response, as well as congruent from incongruent target-locked data were used to evaluate error – correct and incongruent – congruent contrasts.

### Heart-rate metrics

Heart-rate (HR) data was collected through the EEG electrode removed for the tFUS transducer by placing it below the left clavicle. Raw HR data was initially processed manually in MATLAB to ensure correct identification of heart beats, and then all HR metrics were calculated using HRVTool(Vollmer, 2015). We quantified several HR and HRV (heart rate variability) metrics including average HR, R-R interval, standard deviation of the normal-to-normal heartbeat (SDNN), and percentage of successive normal R-R intervals exceeding 50ms (pNN50). A Poincaré plot (RR_n_, RR_n+1_) was constructed to calculate nonlinear measures SD1 (standard deviation perpendicular to the identity line) and SD2 (standard deviation along the identity line), as well as the SD1/SD2 ratio. SD1 is also known as the root mean square of successive R-R interval differences (RMSSD)

### PANAS Mood Ratings

To evaluate mood changed subjects were asked to assess their correct emotional state by completing the Positive Affect and Negative Affect Schedule (PANAS)(Watson et al., 1988). Mood ratings were completed at baseline (prior to stimulation), and again immediately following completion of the experiment. Ratings for positive valence probes (‘interested’, ‘excited’, ‘strong’, ‘enthusiastic’, ‘proud’, ‘alert’, ‘inspired’, ‘determined’, ‘attentive’, ‘active’) were summed to create a PANAS positive score while negative valence probes (‘depressed’, ‘upset’, ‘guilty’, ‘scared’, ‘hostile’, ‘irritable’, ‘ashamed’, ‘nervous,’ ‘afraid’) were summed to create PANAS negative score. Scores were compared with a mixed-factor RM–ANOVA [2 groups × 2 time points (baseline, post-experiment)]

### Behavioral responses

Accuracy and reaction time were measured. Baseline-subtraction was used on RT data to eliminate interference of individual differences. Each subject’ s median baseline RT for congruent and incongruent trials was used (baseline consisted of a simple flanker task on a black background, both groups received sham stimulation). Baseline congruent RT was subtracted from congruent trials (neutral, fear, and oddball), and likewise for incongruent. Groups were then compared using Mann-Whitney U test. Additionally, median RT’s were compared within group using Friedman’s nonparametric test and post-hoc testing with Wilcoxon signed-ranks tests (Bonferroni-corrected).

To access conflict adaption, trials were separated by previous trial congruency (congruent, incongruent), and median congruent RT was subtracted from incongruent to calculated the congruency effect in the RT. These were then compared both across and within group.

### Post error slowing

To assess post error slowing, RTs on fear and neutral trials following an error were compared with median RTs in both congruency conditions using Wilcox signed-rank tests. To assess post error slowing across groups, median RT was subtracted from post error RTs and groups were compared using Mann-Whitney U tests.

### Statistical methods

Statistical analyze were conducted using SPSS Statistics Software SPSS 26.0 (IBM Corporation, Armonk, NY). Only correct responses were considered for analysis. Trials with late RTs, RTs greater than 900ms, or that deviated more than three standard deviations from the individual mean, were excluded from analysis to reduce the effect of outliers.

Both parametric (RM-ANOVA), and non-parametric (between groups: permutation testing, Mann-Whitney U test; within groups: Friedman’s test and Wilcox signed-ranks tests) tests were employed were appropriate and indicated above. Threshold for statistical significance were set at p < 0.05. When necessary normality was confirmed using Kolmogorov-Smirnov tests and Levene’s test for homoscedasticity was used to examine all between group data variance (all p’s > 0.05). All post-hoc tests were Bonferroni-corrected. All data reported is in the format mean ± SEM unless otherwise specified.

## Data Availability

The datasets generated during and/or analyzed during the current study are available from the corresponding author on reasonable request.

## Acknowledgements

We would like to thank Marco Santello of the School of Biological and Health Systems Engineering for the use of his lab and equipment, as well as many thoughtful discussions during these studies.

## Supplemental Materials

### Supplemental Figures

**Figure S1.**
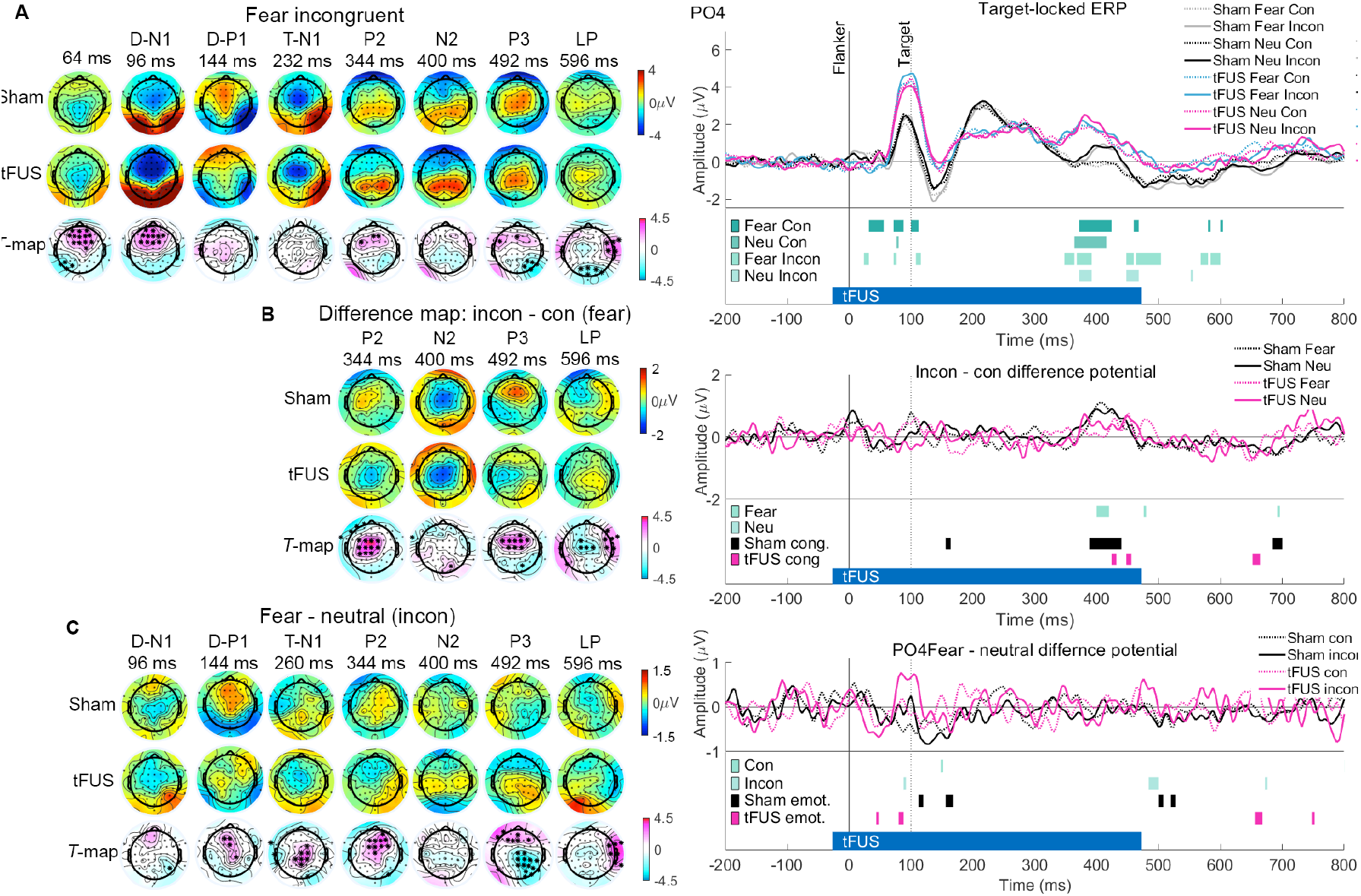
Scalp potentials and ERPs at P4. (A) Scalp-potential voltage topography maps of target locked ERPs displayed in Figure 2. Faces and flanker distractor arrows appeared at 0 ms, and target arrow appeared at 100 ms. tFUS stimulation began 28 ms prior to the faces, and lasted until 472 ms. T value maps plotted below (permutation testing, significant electrodes marked with *; magenta indicates Sham more positive than tFUS, cyan Sham more negative than tFUS. (B) Incongruent minus congruent potential (fear condition) for later ERP peaks in A. (C) Fear – neutral potential (incongruent condition). Right-hand panel displays ERP at PO4 with incon – con and fear – neutral difference potential. Green bars represent permutation testing across groups, magenta and black bars represent main effect of congruency (cong.) or emotion (emot.) within groups (RM-ANOVA). Related to Figure 2, Table S1, Table S2 and Table S3.

**Figure S2.**
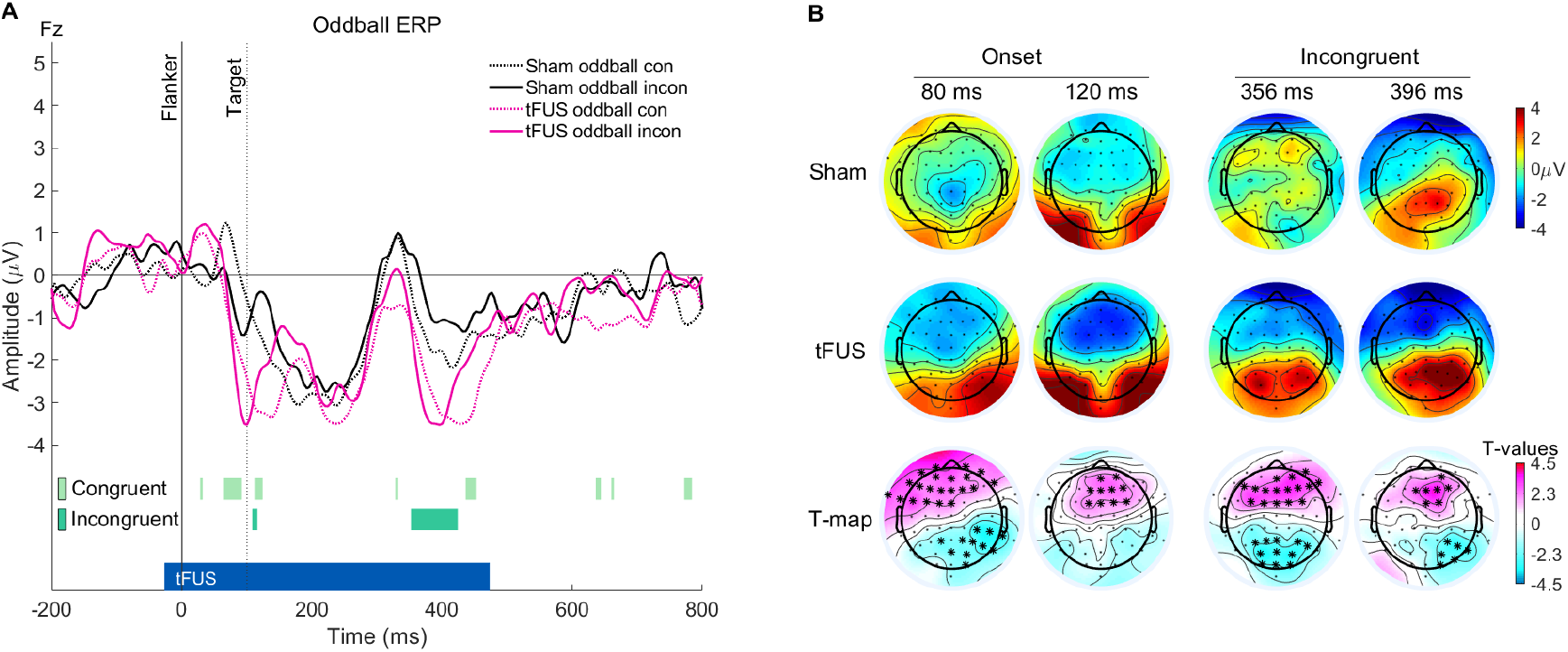
Oddball event related potentials. (A) ERP to the oddball trials at Fz, 0ms represents onset of oddball/scrambled image and distractor flanker arrows, 100 ms marks onset of target arrow. Bars on the bottom of the figure represent the results of Sham vs. tFUS permutation testing (p < 0.05). (B) Scalp maps correspond to significant regions in A. Oddball onset displays the results of pooled incongruent and congruent trials. T values displayed at the bottom of the figure; * represents electrodes p < 0.05 (permutation testing).

### Supplemental Tables

**Table S1.**
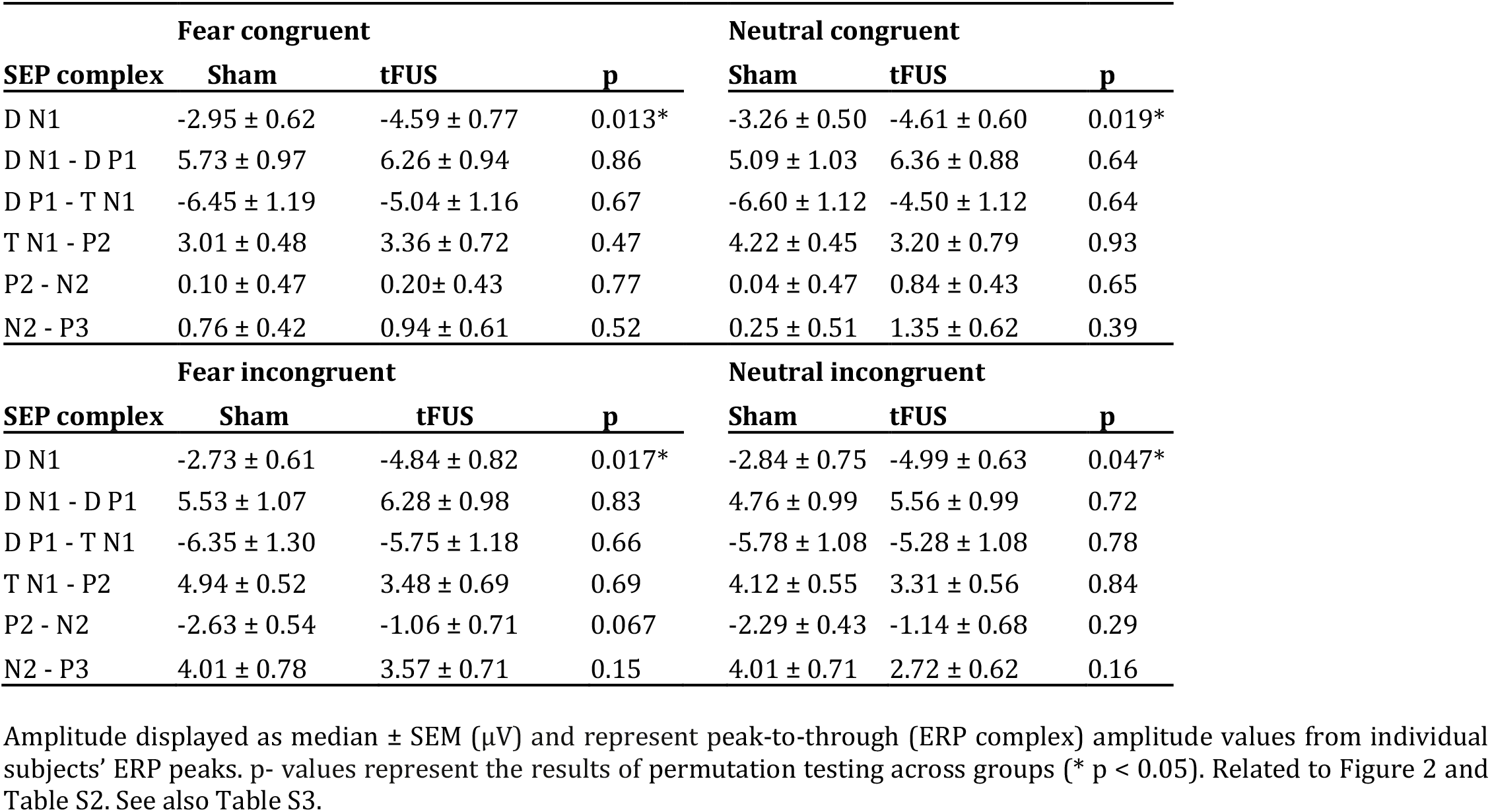
ERP peak-to-peak amplitude at FCz comparing Sham and tFUS groups.

| Fear congruent |  |  |  | Neutral congruent |  |  |
| --- | --- | --- | --- | --- | --- | --- |
| SEP complex | Sham | tFUS | p | Sham | tFUS | p |
| D N1 | -2.95 ± 0.62 | -4.59 ± 0.77 | 0.013* | -3.26 ± 0.50 | -4.61 ± 0.60 | 0.019* |
| D N1 - D P1 | 5.73 ± 0.97 | 6.26 ± 0.94 | 0.86 | 5.09 ± 1.03 | 6.36 ± 0.88 | 0.64 |
| D P1 - T N1 | -6.45 ± 1.19 | -5.04 ± 1.16 | 0.67 | -6.60 ± 1.12 | -4.50 ± 1.12 | 0.64 |
| T N1 - P2 | 3.01 ± 0.48 | 3.36 ± 0.72 | 0.47 | 4.22 ± 0.45 | 3.20 ± 0.79 | 0.93 |
| P2 - N2 | 0.10 ± 0.47 | 0.20 ± 0.43 | 0.77 | 0.04 ± 0.47 | 0.84 ± 0.43 | 0.65 |
| N2 - P3 | 0.76 ± 0.42 | 0.94 ± 0.61 | 0.52 | 0.25 ± 0.51 | 1.35 ± 0.62 | 0.39 |
| Fear incongruent |  |  |  | Neutral incongruent |  |  |
| SEP complex | Sham | tFUS | p | Sham | tFUS | p |
| D N1 | -2.73 ± 0.61 | -4.84 ± 0.82 | 0.017* | -2.84 ± 0.75 | -4.99 ± 0.63 | 0.047* |
| D N1 - D P1 | 5.53 ± 1.07 | 6.28 ± 0.98 | 0.83 | 4.76 ± 0.99 | 5.56 ± 0.99 | 0.72 |
| D P1 - T N1 | -6.35 ± 1.30 | -5.75 ± 1.18 | 0.66 | -5.78 ± 1.08 | -5.28 ± 1.08 | 0.78 |
| T N1 - P2 | 4.94 ± 0.52 | 3.48 ± 0.69 | 0.69 | 4.12 ± 0.55 | 3.31 ± 0.56 | 0.84 |
| P2 - N2 | -2.63 ± 0.54 | -1.06 ± 0.71 | 0.067 | -2.29 ± 0.43 | -1.14 ± 0.68 | 0.29 |
| N2 - P3 | 4.01 ± 0.78 | 3.57 ± 0.71 | 0.15 | 4.01 ± 0.71 | 2.72 ± 0.62 | 0.16 |

**Table S2.**
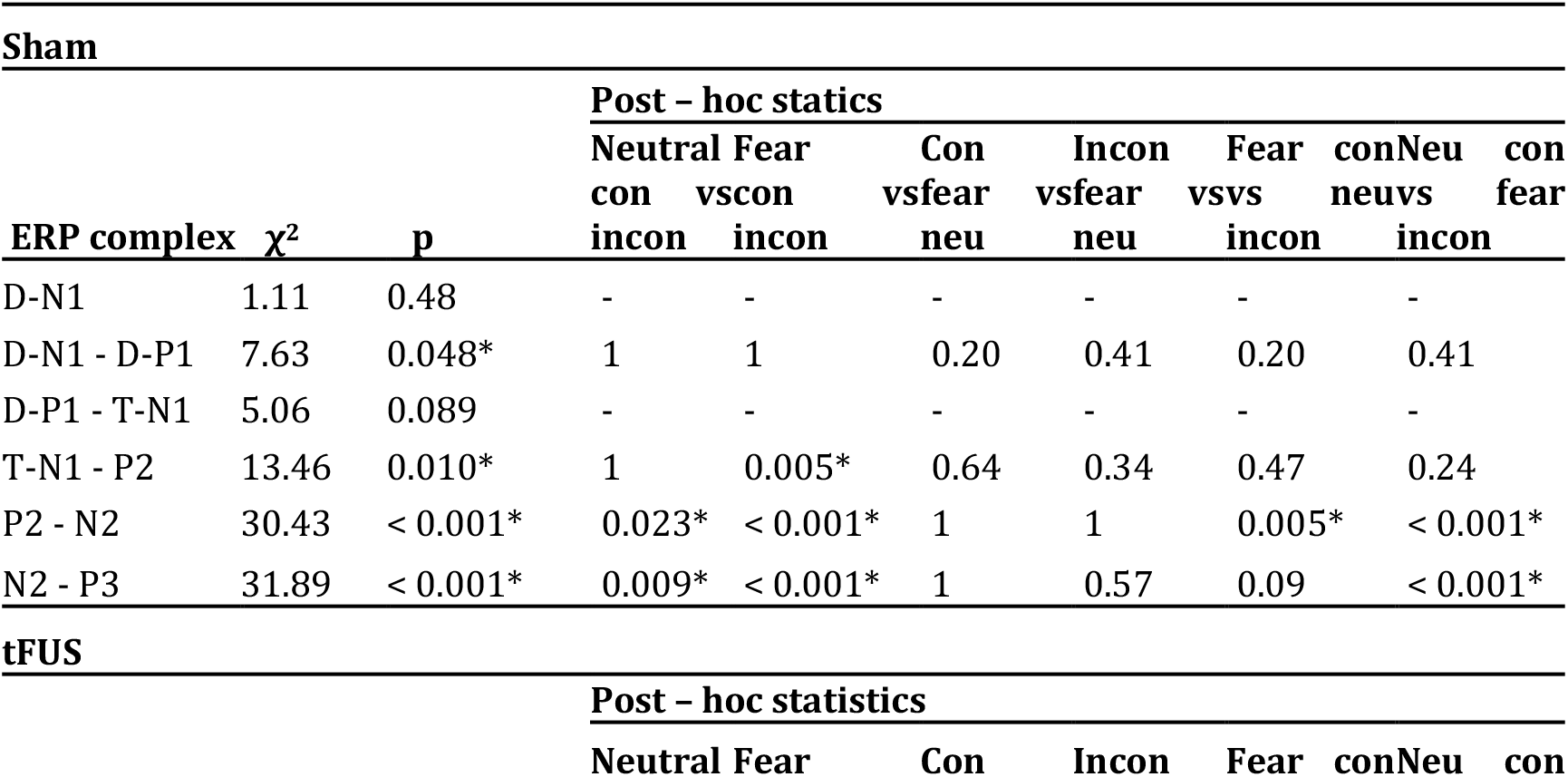
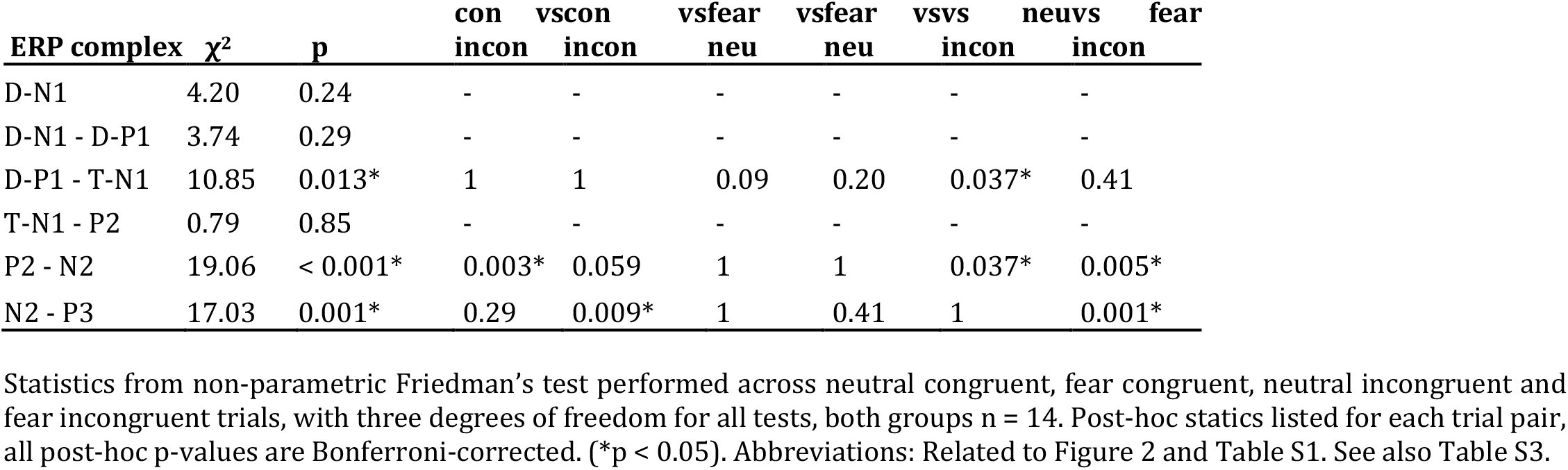
ERP peak-to-peak amplitude at FCz compared within each group using non-parametric Friedman’s test.

**Table S3.** ERP peak latency at FCz comparing Sham and tFUS groups.

| Fear congruent |  |  |  | Neutral congruent |  |  |
| --- | --- | --- | --- | --- | --- | --- |
| SEP peak | Sham | tFUS | p | Sham | tFUS | p |
| D N1 | 108 ± 3 | 104 ± 1 | 0.75 | 104 ± 3 | 100 ± 2 | 0.87 |
| D P1 | 148 ± 3 | 136 ± 3 | 0.14 | 144 ± 4 | 144 ± 4 | 0.80 |
| T N1 | 228 ± 8 | 244 ± 9 | 0.72 | 220 ± 8 | 244 ± 9 | 0.31 |
| P2 | 340 ± 4 | 332 ± 4 | 0.83 | 340 ± 4 | 332 ± 4 | 0.77 |
| N2 | 404 ± 5 | 400 ± 6 | 0.11 | 400 ± 4 | 400 ± 6 | 0.38 |
| P3 | 476 ± 5 | 500 ± 4 | 0.009* | 476 ± 5 | 500 ± 4 | 0.009* |
| Fear incongruent |  |  |  | Neutral incongruent |  |  |
| SEP peak | Sham | tFUS | p | Sham | tFUS | p |
| D N1 | 96 ± 3 | 100 ± 1 | 0.23 | 100 ± 3 | 100 ± 1 | 0.83 |
| D P1 | 144 ± 4 | 140 ± 3 | 0.67 | 144 ± 4 | 144 ± 3 | 0.90 |
| T N1 | 228 ± 8 | 244 ± 8 | 0.95 | 220 ± 7 | 248 ± 8 | 0.10 |
| P2 | 340 ± 5 | 328 ± 3 | 0.32 | 340 ± 7 | 332 ± 4 | 0.22 |
| N2 | 408 ± 9 | 400 ± 5 | 0.95 | 400 ± 8 | 400 ± 5 | 0.84 |
| P3 | 488 ± 5 | 500 ± 4 | 0.077 | 488 ± 5 | 500 ± 4 | 0.095* |

**Table S4.**
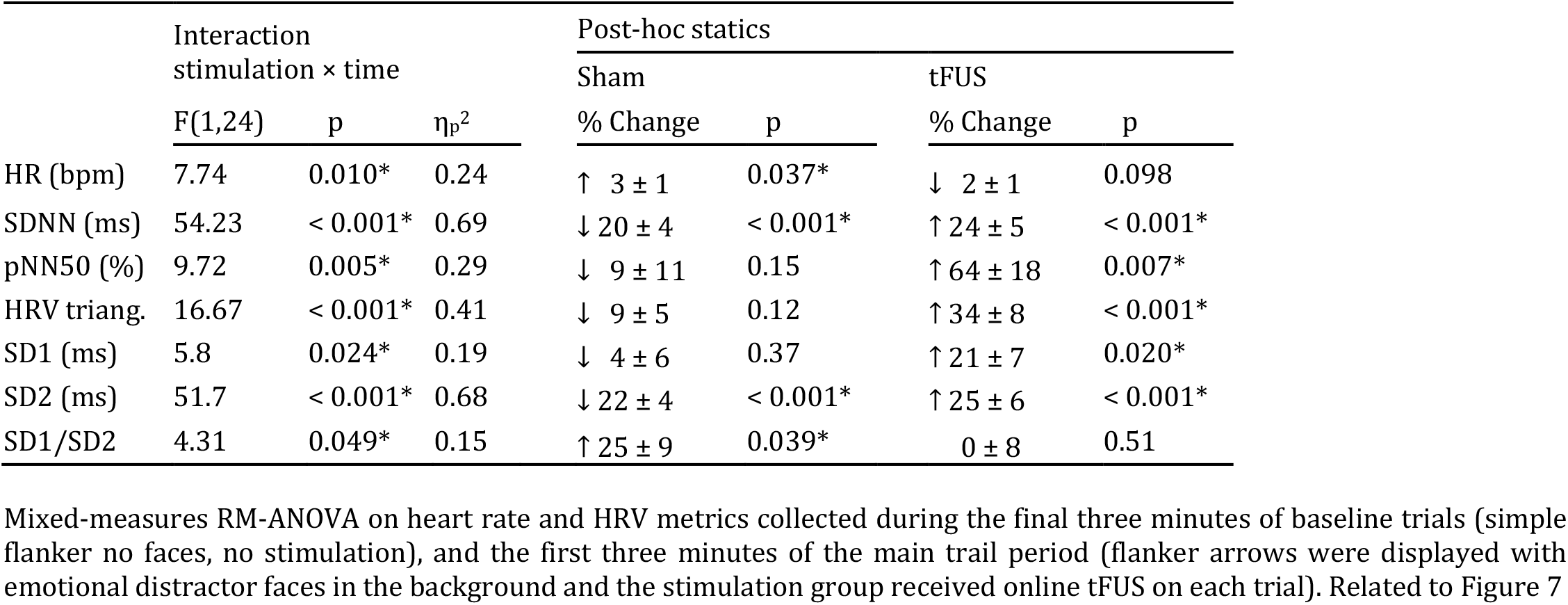
Heart rate and heart rate variability changes differ across groups with emotional faces distractors and tFUS.

**Table S5.**
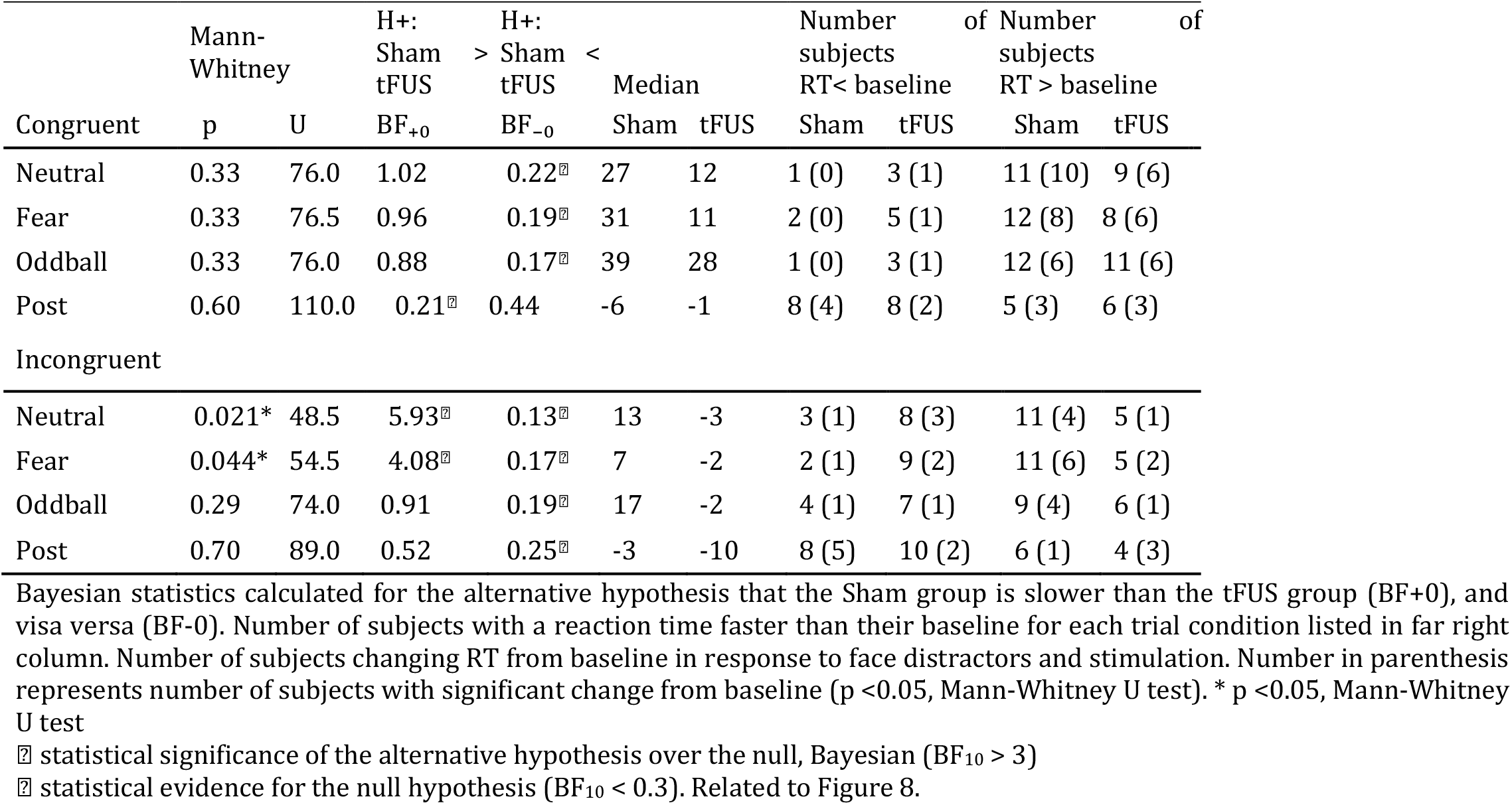
Reaction time group level statistic for baseline-subtracted median RT and change from baseline.

**Table S6.**
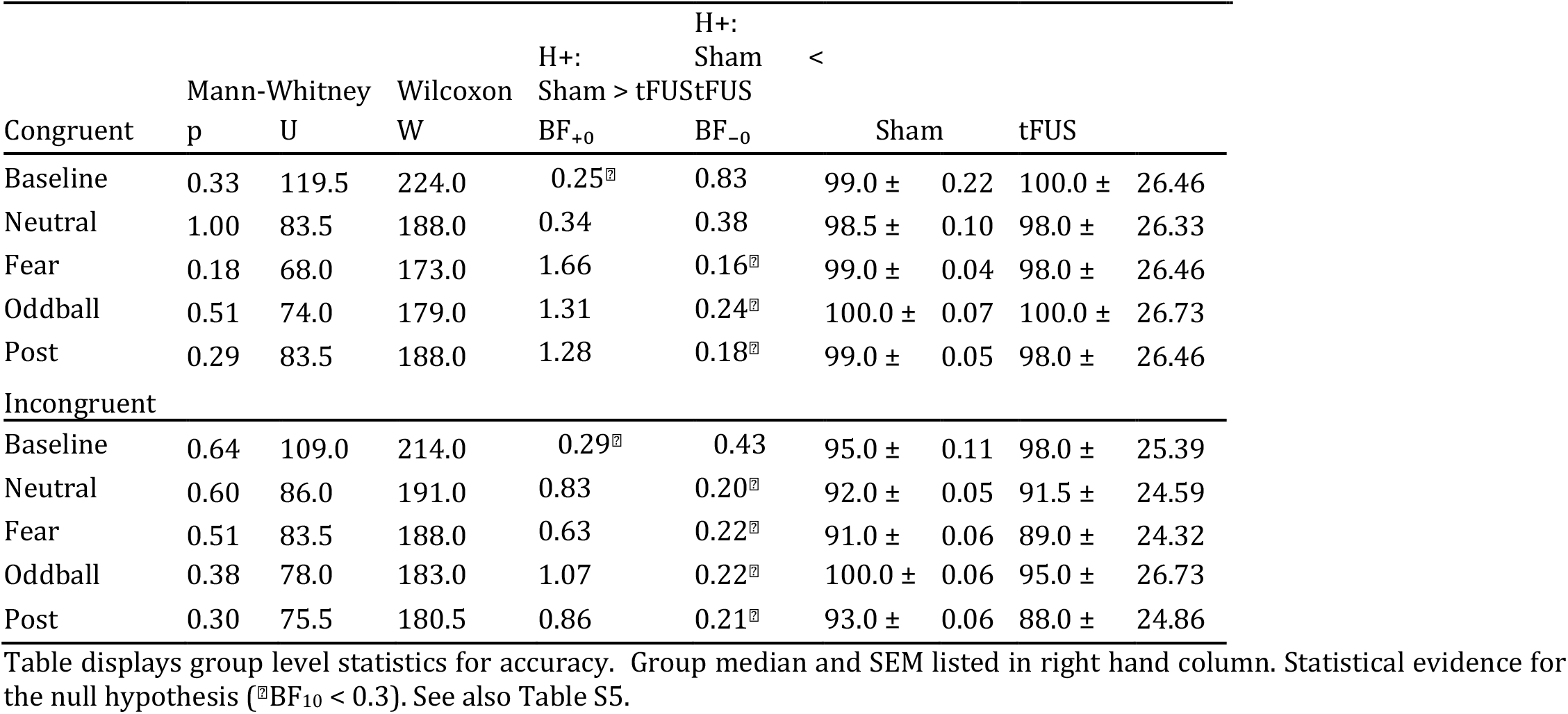
Response accuracy.

